# Targeted tiled amplicon based protocol for sequencing the Hemagglutinin (HA) gene segment of seasonal influenza A and influenza B virus from wastewater at high depth of coverage

**DOI:** 10.1101/2025.10.15.25338105

**Authors:** M.C. Hetherington-Rauth, V. Nguyen, G. Lequia, N. Pysnack, L.A. Bankers, A.E.B. Rossheim, S.R Matzinger

**Affiliations:** Colorado Department of Public Health & Environment

## Abstract

Wastewater based epidemiology has emerged as a compelling tool to monitor the spread and evolution of pathogens of public health concern. Next generation sequencing (NGS) of pathogens detected in wastewater enables sequence characterization which is essential for monitoring the changing genomic landscape of pathogens. Influenza virus, with its potential to cause epidemics and pandemics, poses a serious risk to human health. Genomic surveillance of the virus is essential for safeguarding public health by monitoring the virus’s evolution, and developing preventive vaccines and therapeutics. As such, methods for successfully sequencing influenza virus at a consistent high depth of coverage across the genome can aid in this endeavor. Here, we present a novel targeted tiled amplicon based sequencing protocol that uses short tiled amplicons (<250 bp in length) to successfully capture the Hemagglutinin (HA) gene segment of seasonal influenza A subtypes (H1 and H3) and Influenza B at high depth of coverage. We observed near consistent coverage across the HA gene segment for wastewater samples that had influenza viral target dPCR detections of at least 10^3^ copies/L. We were able to successfully detect low frequency single nucleotide variants (SNVs) at high depth of coverage demonstrating the utility of the data to characterize the diversity of circulating influenza A and B viruses at the community level. Our approach is flexible and future directions include expanding this approach to sequence additional influenza virus HA subtypes and gene segments.

## Introduction

Influenza virus causes acute viral respiratory disease in humans and results in approximately 3-5 million severe cases and 290,000 to 650,000 deaths worldwide each year (*1*). In the United States, influenza results in 140,000 to 710,000 hospitalizations and 12,000 to 52,000 deaths per year, and places a high burden on the health care system (*2*). Although vaccines and therapeutics exist for the prevention and treatment of influenza, public health concern remains due to the virus’s pandemic potential (*3*).

Influenza virus exhibits high genetic diversity. The influenza A and B virus genomes are composed of 8 negative-sense RNA gene segments which can result in genetic reassortment of individual gene segments resulting in antigenic shift (*4*, *5*). Additionally, influenza viruses replicate with low fidelity, resulting in a high mutation rate (*6*). Together, genetic reassortment and low replication fidelity contribute to the emergence of new strains that can evade immune system detection and/or result in the introduction of highly pathogenic strains. Influenza A virus subtypes H1N1 (InfA H1N1) and H3N2 (InfA H3N2), and influenza B virus cause annual seasonal epidemics. However, many additional avian influenza A virus subtypes exist including H5N1, H7N9, and H9N2, which have the potential to spillover into humans (*7*).

Traditional clinical surveillance systems, where samples taken from patients are tested and sequenced, are often limited to symptomatic and/or severe cases from patients that sought medical care and/or were hospitalized. These cases represent only a small fraction of disease transmission and are often subject to biases due to healthcare disparities among poor and underserved communities (*8*). Wastewater based surveillance methods, which include both PCR-based detection and quantification methods, and next generation sequencing (NGS), expand upon traditional surveillance methods by capturing asymptomatic and less severe cases as well as cases from underserved communities with limited clinical testing capacity and/or utilization (*9*, *10*). Additionally, wastewater surveillance can be cost effective relative to traditional surveillance systems, since a single wastewater sample represents an aggregated community level sample.

Wastewater detection and quantification methods, which include RT-qPCR (reverse transcription-quantification PCR), ddPCR (digital droplet PCR), and dPCR (digital PCR), are often designed off of short, highly specific, and well-conserved regions of the genome, and thus are limited in their ability for genomic characterization (*11*). Such methods have been applied to successfully detect and quantify influenza virus in wastewater and monitor trends of influenza in communities (*12*, *13*). Next generation sequencing (NGS) of wastewater samples, however, allows for the sequence characterization of specific genetic regions or the entire genome.

Sequence characterization can be used to identify single nucleotide variants (SNVs), monitor clade abundances, and track the virus’s evolution, which can inform public health decision-making and the development of vaccines and therapeutics. During the COVID-19 pandemic, wastewater NGS surveillance was used to monitor variant abundances in communities, which corroborated trends in variant estimations in clinical surveillance systems (*14–18*). Additionally, wastewater NGS surveillance allowed for the early detection of new variants introduced into communities before being detected in clinical based surveillance systems (*19–22*).

The success of sequencing SARS-CoV-2 from wastewater highlights the potential of sequencing additional viral pathogens from wastewater. Previous studies have demonstrated the feasibility of sequencing influenza from wastewater using targeted amplicon, tiled amplicon, and hybrid capture methods (*12*, *23–27*). These studies also highlight the challenge of sequencing influenza viruses from wastewater at consistent and high coverage across individual gene segments including high consequence gene segments, such as HA, especially at lower levels of influenza virus concentrations.

In this paper, we present a targeted tiled amplicon primer scheme to sequence the HA gene segment of seasonal Influenza A H1 (InfA H1), Influenza A H3 (InfA H3), and Influenza B (InfB) from wastewater. We focused our efforts on sequencing the HA gene segment because of the role its antigenic structure plays in immune escape and vaccine development (*28–31*). We used the program Olivar to design a tiled amplicon primer scheme that accounts for the known diversity of influenza virus circulating in the human population. We demonstrate that this targeted amplification strategy is sensitive enough to sequence portions of the HA gene segment at wastewater influenza virus concentrations as low as 10^2^ copies/L, but performs best at influenza virus concentrations of 10^3^ copies/L or higher. Our approach can be expanded to sequence other influenza virus subtypes and gene segments from wastewater, including avian influenza A viruses capable of severe human infection, making it a valuable tool for public health surveillance.

## Results

### Tiled Amplicon Primer Scheme

We designed three primer schemes that amplify the HA gene segment for InfA H1, InfA H3 and InfB, respectively. The HA gene segment primer scheme for 1) InfA H1 captured 90.3% of the HA gene segment and consisted of 12 primer pairs with an average amplicon size of 215 basepairs, 2) InfA H3 captured 86.4% of the HA gene segment and consisted of 13 primer pairs with an average amplicon size of 196 bps, and 3) InfB captured 85.1% of the HA gene segment and consisted of 13 primer pairs with an average amplicon size of 193 bps (**Tables 1, S1-S3**).

**Table 1.** Tiled amplicon primer scheme metrics.

| *Influenza Subtype* | % Coverage of the HA Gene Segment Included | Number of Primer Pools | Number of Amplicons | Number of Primers | Average Amplicon Size (bps) | Average Primer Size (bps) |
| --- | --- | --- | --- | --- | --- | --- |
| InfA H1 | 90.3 | 2 | 12 | 24 | 214 | 25 |
| InfA H3 | 86.4 | 2 | 13 | 26 | 196 | 24 |
| InfB | 85.1 | 2 | 13 | 26 | 192 | 23 |

### HA Gene Segment Assembly

*Positive controls.* We sequenced whole virus controls in both a background of nucleus free water (NFW) and RNA extracted pooled wastewater that tested negative for influenza A and influenza B by dPCR. Whole virus controls followed an eight step 10-fold serial dilution. Depth of coverage and percent coverage was similar between controls in NFW and RNA extracted pooled wastewater backgrounds at high concentrations (**Tables S4-S6**). Depth of coverage and percent coverage was lower for controls in RNA extracted pooled wastewater background than for controls in NFW background at lower concentrations (**Fig S1**).

*Clinical samples.* We sequenced clinical samples collected from January 2025 to March 2025 to determine if our primer scheme was robust to amplicon drop outs against the current influenza diversity observed in circulation during the 2024-2025 respiratory season. We observed complete coverage of the HA gene segment covered by our primer scheme and average depths of coverage > 50,000 reads for InfA H1, InfA H3 and InfB (**Table 2**). We observed near consistent depth of coverage across the HA gene segment for InfA H1 and InfA H3 (**Fig 1 A, B**). We observed a slight drop in depth of coverage across amplicons 8 and 9 for the InfB primer scheme (**Fig 1 C**). Upon manual inspection, we observed one mutation occurring at >97% frequency in both of the reverse primers of amplicon 8 and 9 for all clinical samples; however, further investigation is needed to determine if the drop in coverage can be explained due to primer sequence mismatches.

**Fig. 1.**
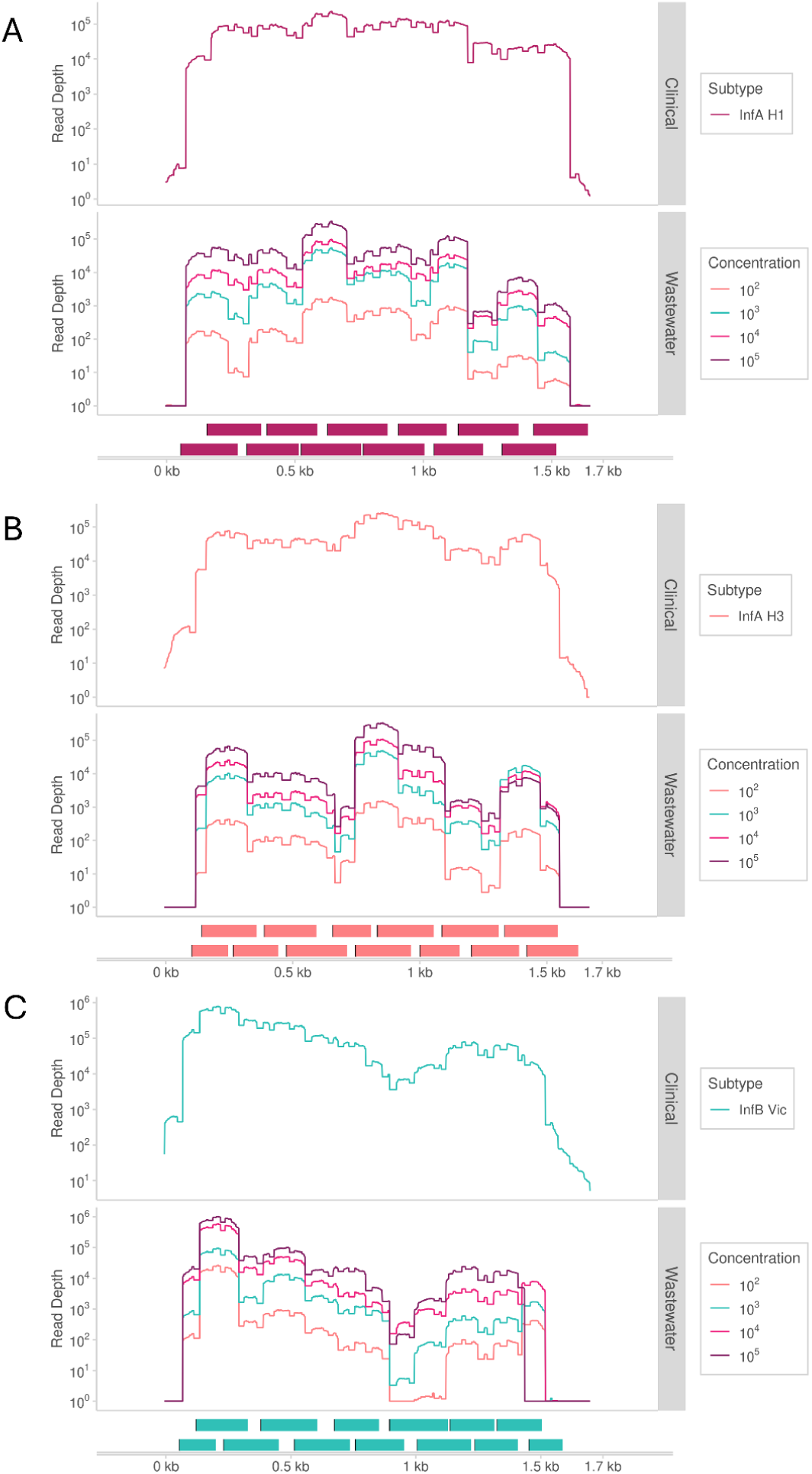
Depth of coverage across the HA gene segment. Panels show (A) InfA H1, (B) InfA H3, and (C) InfB. For each panel, the top track shows the average depth of coverage for clinical samples. The middle track shows the average depth of coverage for wastewater samples. Colors show the concentration of influenza virus (copies/L) present in the wastewater sample grouped into four bins (10^2^, 10^3^, 10^4^ and 10^5^ copies/L). The bottom track shows the tiled amplicon scheme. Read depth is shown in log scale.

**Table 2.**
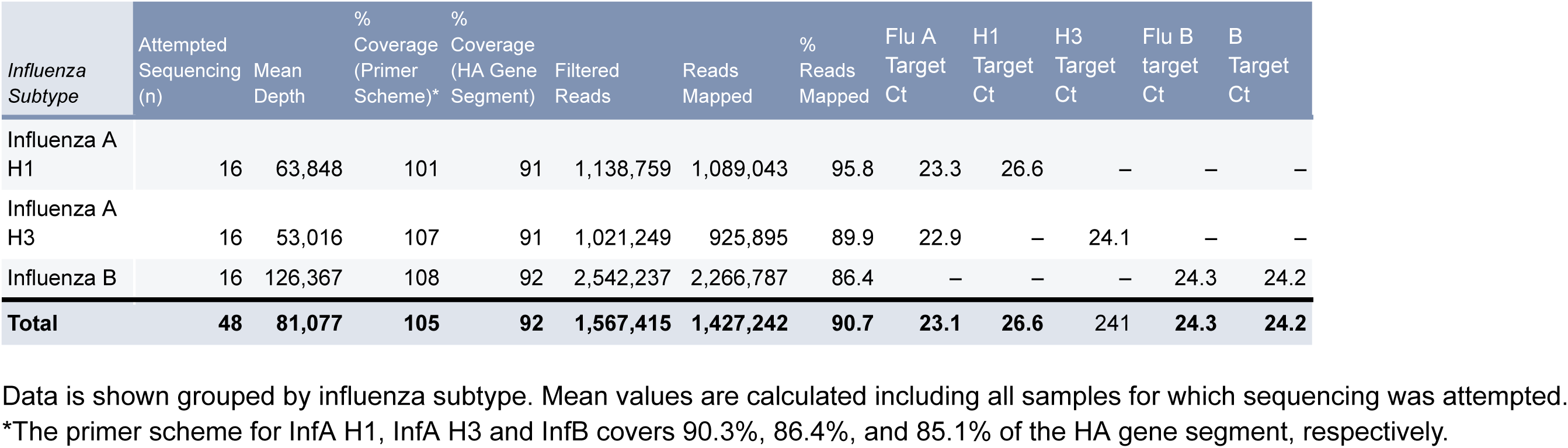
Assembly metrics of clinical samples.

*Wastewater samples.* We sequenced the HA gene segment on a total of 188 InfA and InfB positive wastewater samples (60 InfA H1, 59 InfA H3, and 69 InfB) collected during the 2024-2025 respiratory season. We binned samples into four groups corresponding to influenza virus detection concentrations of 10^2^, 10^3^, 10^4^ and 10^5^ copies/L (**Table 3**). In general, we observed that the percent coverage and mean depth of coverage increased with increasing virus concentration (**Fig 2**).

**Fig. 2.**
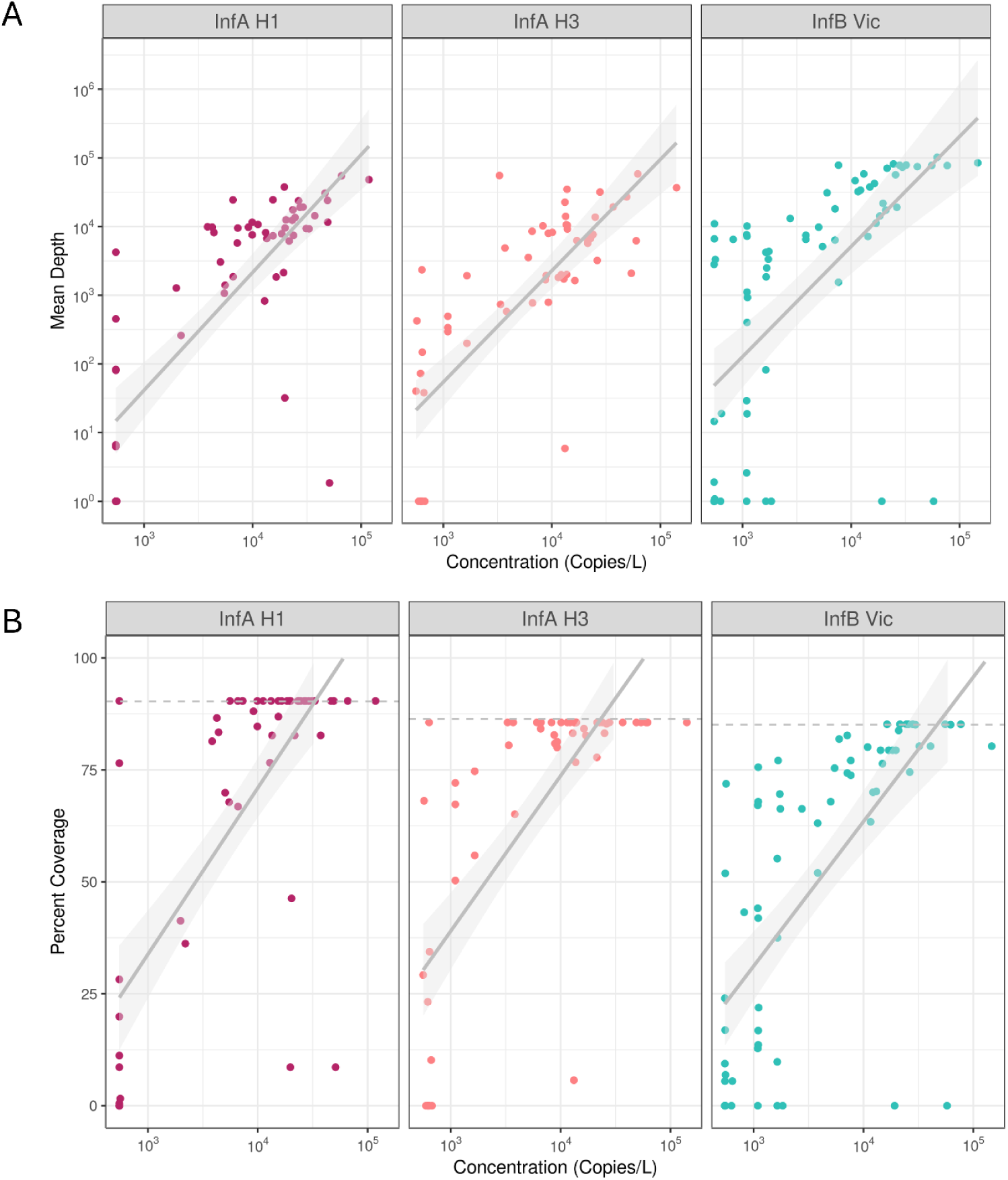
Effect of Influenza virus concentration in wastewater samples on mean depth of coverage and percent coverage of the HA gene segment. Scatter plot showing the relationship between the concentration (copies/L) of influenza virus detected in wastewater and (A) the mean depth of coverage and (B) the percent coverage of the HA gene segment. Both the mean depth of coverage and the percent coverage of the HA gene segment increased with increasing Influenza virus concentration. In (B) the percent of the HA gene segment covered by the primer scheme is indicated by the horizontal dashed line. The primer scheme for InfA H1, InfA H3 and InfB covers 90.3%, 86.4%, and 85.1% of the HA gene segment, respectively. The linear regression with 95% CI interval is shown in grey. Concentration and mean depth of coverage are shown in log scale.

**Table 3.** Assembly metrics of wastewater samples grouped by concentration.

| *Influenza Subtype* | *Wastewater Concentration Group* | Attempted Sequencing (n) | Mean Depth | % Coverage (Primer Scheme)* | % Coverage (HA Gene Segment) | Filtered Reads | Reads Mapped | % Reads Mapped | Wastewater Concentration (Copies/L)<sup>†</sup> |
| --- | --- | --- | --- | --- | --- | --- | --- | --- | --- |
| Influenza A H1 | 10<sup>2</sup> | 15 | 324 | 17 | 16 | 1,730,834 | 5,783 | 0.3 | 549 |
|  | 10<sup>3</sup> | 15 | 7,027 | 85 | 77 | 1,713,415 | 127,834 | 7.3 | 5,963 |
|  | 10<sup>4</sup> | 29 | 13,904 | 91 | 82 | 1,390,059 | 254,768 | 18.3 | 26,207 |
|  | 10<sup>5</sup> | 1 | 48,235 | 100 | 90 | 1,361,062 | 909,246 | 66.8 | 118,000 |
| Subtotal |  | 60 | 9,362 | 71 | 64 | 1,555,608 | 171,696 | 11.8 | 19,121 |
| Influenza A H3 | 10<sup>2</sup> | 14 | 219 | 21 | 18 | 673,898 | 4,155 | 0.5 | 614 |
|  | 10<sup>3</sup> | 17 | 5,903 | 90 | 77 | 842,299 | 111,752 | 9.5 | 4,973 |
|  | 10<sup>4</sup> | 26 | 12062 | 95 | 82 | 961,591 | 225,974 | 22.1 | 23,800 |
|  | 10<sup>5</sup> | 1 | 36,837 | 100 | 86 | 1,058,432 | 670,526 | 63.4 | 141,000 |
| Subtotal |  | 58 | 7825 | 76 | 65 | 858,853 | 146,617 | 13.9 | 14,708 |
| Influenza B | 10<sup>2</sup> | 16 | 1,892 | 17 | 15 | 1,022,374 | 38,418 | 3.4 | 606 |
|  | 10<sup>3</sup> | 28 | 7,902 | 58 | 50 | 1,045,063 | 157,063 | 11.8 | 3,116 |
|  | 10<sup>4</sup> | 24 | 46,366 | 86 | 73 | 1,979,942 | 926,574 | 43.3 | 26,945 |
|  | 10<sup>5</sup> | 1 | 84,685 | 94 | 80 | 2,393,578 | 1,591,955 | 66.5 | 147,000 |
| Subtotal |  | 69 | 21,000 | 59 | 50 | 1,384,521 | 418,003 | 21.6 | 15,723 |
| **Total** |  | **187** | **13180** | **68** | **59** | **1,276,374** | **254,801** | **16.1** | **14,727** |

Data is shown grouped by influenza subtype and wastewater concentration group. Mean values are calculated including all samples for which sequencing was attempted.
\*The primer scheme for InfA H1, InfA H3 and InfB covers 90.3%, 86.4%, and 85.1% of the HA gene segment, respectively.
<sup>†</sup>Concentration corresponds to that of the InfA H1 dPCR target for InfA H1 subtype, InfA H3 dPCR target for InfA H3 subtype and InfB dPCR target for InfB.

We observed near complete coverage of the HA gene segment covered by the primer scheme at 10^3^ copies/L and higher with a noticeable increase in percent coverage for InfA H1 and InfA H3 from 10^3^ to 10^5^ copies/L (**Table 3**). For both InfA H1 and InfA H3, percent coverage was more variable in samples at 10^2^ copies/L (SD = 32.0 for InfA H1 and SD = 32.6 for InfA H3) compared to samples at 10^3^ (SD = 19.6 for InfA H1 and SD = 12.8 for InfA H3) and 10^4^ (SD = 24.4 for InfA H1 and SD = 18.3 for InfA H3) copies/L. The depth of coverage across the InfA H1 HA gene segment was consistent except for a noticeable drop in coverage in the regions corresponding to amplicons 10 and 12. The depth of coverage across the InfA H3 HA gene segment was relatively consistent.

For InfB, we observed near complete coverage of the HA gene segment covered by the primer scheme at 10^4^ copies/L and higher and a noticeable increase in percent coverage from 10^3^ to 10^4^ copies/L and (**Table 3**). The lower percent coverage observed for InfB at 10^2^ and 10^3^ copies/L compared to that of InfA H1 and InfA H3 is likely due to the drop in coverage we observed across the region corresponding to amplicons 8 and 9 that was also observed in clinical samples. The variability in percent coverage was relatively consistent across concentrations (SD = 25.9 for samples at 10^2^ copies/L, SD = 33.5 for samples at 10^3^copies/L, and SD = 27.4 for samples at 10^4^ copies/L) for InfB.

We performed reciprocal mapping by aligning reads from InfA H1, InfA H3 and InfB to the other two reference sequences in each case, in order to assess the specificity of the InfA H1, InfA H3 and InfB primer scheme for its respective targeted subtype or lineage. In all cases we observed no more than two reads aligning to non-targeted reference sequences. This was true even among wastewater samples that were positive for all targets (InfA H1, InfA H3 and InfB) using dPCR.

### Single Nucleotide Variants (SNVs)

We performed variant calling against the 2023-2024 influenza virus vaccine strain sequences to determine the number of and frequency of SNVs occurring in wastewater samples (**Fig 3**).

**Fig. 3.**
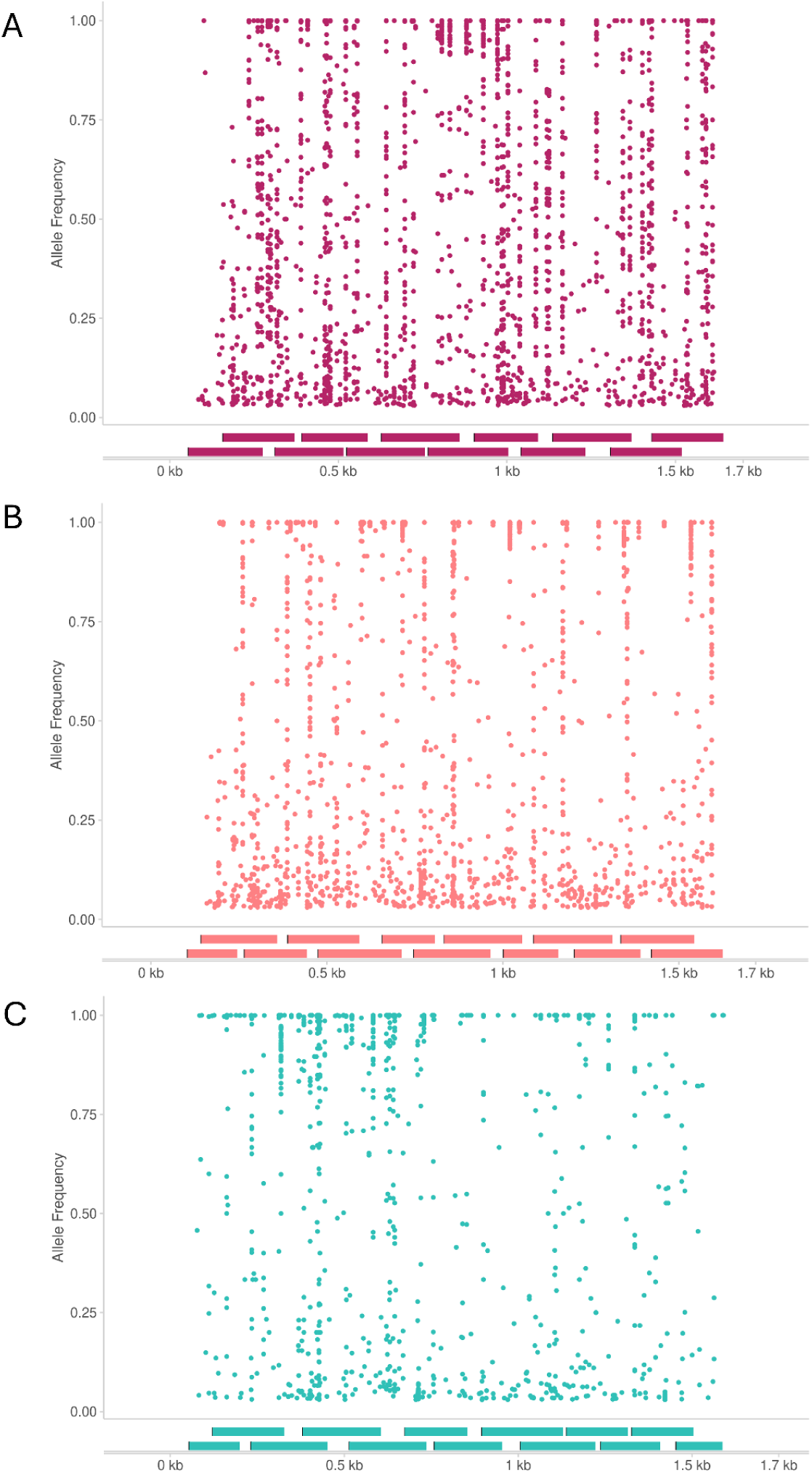
Allele frequencies of single nucleotide variants (SNVs) detected in wastewater. The allele frequencies of single nucleotide variants (SNVs) relative to the vaccine strain detected in wastewater samples across the HA gene segment for (A) InfA H1, (B) InfA H3 and (c) InfB. Mixed proportions of alternative and vaccine allele frequencies were observed, indicative of a mixed sample, as is expected for wastewater samples. The tiled amplicon scheme is shown below each graph.

Mixed frequencies of SNVs are expected in wastewater samples and are indicative of the diversity and proportion of circulating influenza viruses in a community. We detected a total of 343, 389, and 291 unique SNVs among all InfA H1, InfA H3, and InfB wastewater samples relative to the vaccine reference sequences, respectively. Of those, 333, 377, 258 SNVs occurred at mixed frequencies between 0.03 and 0.97 for InfA H1, InfA H3, and InfB, respectively. The average read depth for SNVs occurring at frequencies near our detection cut off between 0.03 and 0.05, was >100x, >500x and >800x for wastewater samples with influenza virus concentrations at 10^3^, 10^4^ and 10^5^ copies/L, respectively (**Fig 4**).

**Fig. 4.**
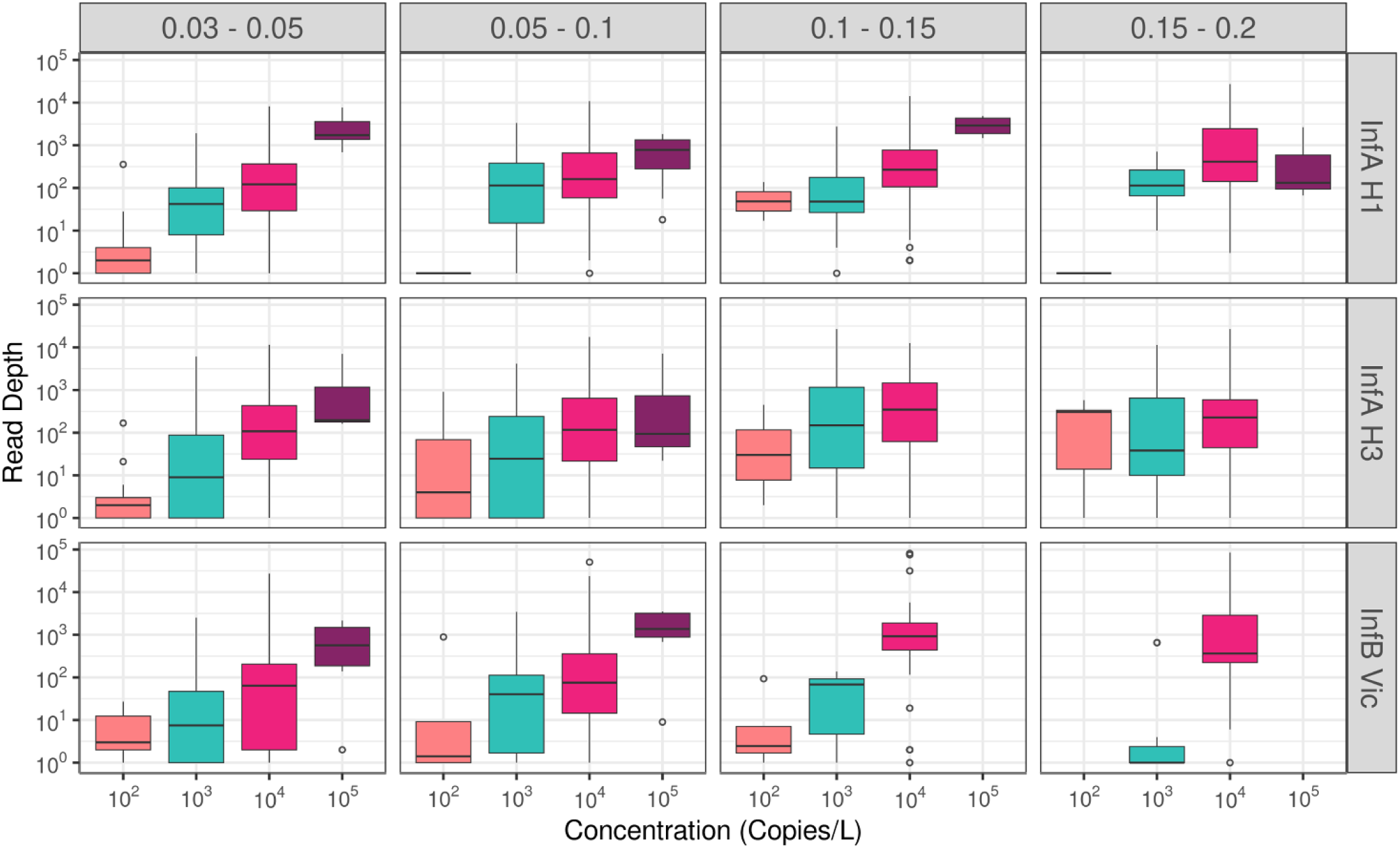
Depth of coverage for the detection of low frequency SNVs. The depth of coverage for SNVs detected from 0.03 to 0.2 frequency is shown in relation to the concentration of influenza (in copies/L) detected in wastewater samples for each InfA H1, InfA H3 and InfB. We binned SNV frequency into four bins (0.03 - 0.05, 0.05 - 0.1, 0.1 - 0.15, 0.15 - 0.2) as shown along the top of the figure. The concentration of influenza detected in wastewater is grouped into 4 bins (10^2^, 10^3^, 10^4^ and 10^5^) and is shown along the x-axis. The minimum frequency threshold for calling SNVs was set to 0.03. Read depth is shown in log scale.

## Discussion

Wastewater based pathogen surveillance has emerged as a promising tool for enhancing the surveillance of pathogens of public health concern (*9*, *32–34*). Sequencing influenza virus from wastewater has presented challenges due to the fragility of viral RNA and the virus’s high genomic diversity and segmented genome. Here, we present a tiled amplicon primer scheme and an optimized sequencing method that allows for high depth of coverage across the HA gene segment of InfA H1, InfA H3, InfB virus.

Wastewater is a complex matrix that negatively impacts the stability of viral RNA, especially that of enveloped viruses, such as influenza, and other respiratory pathogens including coronaviruses and respiratory syncytial virus (RSV) (*34*, *35*). Enveloped viruses have a labile lipid envelope, which makes them more susceptible to environmental conditions including pH, temperature, surfactants, and other microorganisms (*34*, *36*). As such, viral RNA in wastewater is expected to be highly fragmented. This creates a challenge for sequencing, especially methods that require intact viral RNA in high concentrations.

Given that viral RNA in wastewater is expected to be fragmented, methods that focus on short, targeted regions, or use short amplicons across a larger genomic region, are expected to yield more consistent coverage than approaches that attempt to capture large genomic regions. For example, attempts at sequencing short targeted gene regions ranging in size from 100 bp to 1000 bp using nested primers have proven successful for confirming detection of influenza virus in wastewater (*24*, *25*). Given this success, tiled amplicon approaches that capture multiple short regions across a gene segment prove promising. For example, John et al. (*27*) developed a tiled amplicon primer panel, with an amplicon size of ∼400 bps, that captured the HA, NA, and M gene segments of influenza A from wastewater. Although John and colleagues observed drop out regions, this was likely due to the diversity of influenza causing primer mismatches rather than the inability to capture fragmented viral RNA.

Building upon these methods, we designed a tiled amplicon primer scheme that amplifies short regions, <250 bp amplicons, of the HA gene segment. With our primer scheme, we observed relatively consistent coverage across the entire gene segment. Taken together with other studies, our results highlight that approaches for targeted sequencing of influenza viral RNA from wastewater, and more generally, targeted sequencing of viral RNA from other enveloped viral pathogens, should focus on amplifying shorter regions across the genome rather than attempting to amplify full gene segments or large genomic regions with only a small number of primer pairs.

In addition to the complex matrix of wastewater, sequencing influenza from wastewater is further complicated by the virus’s high genomic diversity. The genomes of influenza A and B are composed of 8 gene segments. The HA and NA gene segments of influenza A have 16 (H1-H16) and 9 (N1-N9) subtypes that are commonly found in avian species, respectively (*2*). In wastewater, it is possible to find multiple subtypes, including avian subtypes, such that wastewater is essentially a soup of influenza subtypes and gene segments. Furthermore, high levels of genetic diversity are present within subtypes and across host species, increasing the probability that targeted approaches may either experience drop out regions or not capture the full breadth of genetic diversity present in a sample. Several indiscriminate or agnostic approaches have been implemented that attempt to capture the complete diversity of the influenza virus, including a universal primer scheme (*23*) and hybrid capture methods (*12*, *26*).

Despite having high genetic diversity, influenza A virus exhibits highly conserved regions at the 3’ and 5’ termini of each gene segment which are important regions for transcription and replication of the viral genome. A universal primer scheme, consisting of only 3 primer pairs that simultaneously amplifies all 8 segments and all influenza A subtypes, was created by taking advantage of these highly conserved regions (*37–39*). Similar universal primers exist for influenza B (*40*). This approach is widely used to sequence influenza viral RNA from clinical samples collected from infected patients. Such a universal method would be ideal for sequencing influenza viral RNA from wastewater when there is no prior knowledge of what influenza A viral subtypes are expected or when the goal of the study is to describe what is present. However, applying this universal primer scheme to wastewater has proved challenging due to the fragmented nature of influenza viral RNA in wastewater. Because the universal primer pairs sit at the ends of each gene segment, unfragmented gene segments will amplify with more consistency.

A hybrid capture approach, such as that used in respiratory sequencing panels, offers a solution to capture a wide breadth of influenza diversity. Xiao et al. (*41*) designed a panel using the complete diversity of influenza A, B and C. Although the panel was not designed specifically for wastewater samples, it was designed in a conscious effort to capture fragmented viral RNA, and could be applied to this sample type. Other respiratory and viral pathogen panels have been utilized to track influenza virus, identify specific strains, and corroborate clinical case data (*12*, *26*, *42*).

While hybrid capture panels can effectively target genomic diversity within and between viral pathogens, they often sacrifice percent coverage and depth of coverage, especially as more targets are added to the panel (*43*). High depth of coverage is required to confidently call single nucleotide variants (SNVs) at low frequencies and perform deconvolution to estimate clade and lineage abundances. For example, Grubauagh et al. (*44*) (2019) recommend sequencing to a depth of at least 400x to detect intrahost SNVs occurring at 0.03 frequency given 1,000 virus copies. Additionally, Freyja, a popular software for performing deconvolution, uses a depth-weighted least absolution deviation regression to estimate abundance such that it gives priority to sites as a function of their sequencing depth (*14*).

In contrast to hybrid capture or universal primer methods, our targeted tiled amplicon primer scheme presented here, although limited and specific to the HA gene segments of seasonal influenza A (H1N1 and H3N2) and influenza B, requires only 12 or 13 primer pairs to capture the near complete HA segment with consistent coverage at mean sequencing depths > 5,000x for samples with influenza detected with at least 10^3^ copies/L. The increased depth of coverage gained by this targeted approach increases the sensitivity to detect SNVs present at low frequencies, which is necessary to surveil for mutations that may indicate increased virulence, drug resistance, and vaccine escape. Part of this strength stems from our use of Olivar (*45*), a tool developed to design primer schemes that account for the known genomic diversity of influenza. In this way, the primer scheme was designed to avoid genetic regions of known high diversity and thus increase robustness to amplicon dropouts caused by mismatches between the primer sequences and circulating viral variants. This targeted sequencing approach is flexible enough that it could be adapted to additional HA subtypes, including avian influenza A viruses as well as other gene segments that affect therapeutic effectiveness (46) and contribute to virulence (*47*).

Ultimately, the tradeoff between capturing all genomic diversity and achieving high sequencing depth reflects what Singer et al. (*33*) describes as targeting ‘known-knowns,’ ‘known-unknowns,’ and ‘unknown-unknowns.’ That is, should priority be given to targeted methods that generate high depth of coverage to surveil for mutations that could indicate increased virulence, drug resistance, and vaccine escape, or should priority be given to hybrid capture and even metagenomic approaches that may be able to detect increases in known pathogens of concern (e.g. highly pathogenic avian influenza) and even unknown emerging novel strains. Both are important questions directed towards public health surveillance and likely require different approaches that should be used in parallel.

## Conclusions

In summary, we present a targeted tiled amplicon primer scheme that captures the seasonal diversity of the HA gene segment of influenza A H1 and H3 and influenza B from wastewater samples at concentrations as low as 10^2^ copies/L. This targeted approach provides high depths of coverage (>5,000x) that are relatively consistent across the length of the gene segment for wastewater samples with influenza concentrations detected at 10^3^ copies/L or higher. We show that with this targeted sequencing method, SNVs present at frequencies as low as 3% are supported by high read depths (>100x) for wastewater samples with influenza virus concentrations at 10^3^ copies/L or higher. In addition to expanding our primer scheme to additional gene segments, future efforts could focus on optimizing the wet bench protocol to enable a multiplex reaction, allowing all primer schemes to be combined into a single workflow. A multiplex reaction would improve efficiency by reducing lab time and reducing the consumption of reagents. As influenza continues to be a threat to human health with pandemic potential, performing wastewater sequencing surveillance alongside traditional surveillance methods, can aid in monitoring the virus’s evolution and emerging variants of concern and contribute towards vaccine development and therapeutics.

## Materials and Methods

### Wastewater sample selection

We selected samples collected from wastewater utilities within Colorado’s sentinel surveillance system between August 2024 and January 2025. Based on sample availability, we selected a representative subset of wastewater samples that were tested for the presence and quantification of Influenza HA H1 and H3 and Influenza B gene copies using dPCR (as described below) irrespective of wastewater utility or sampling location. The selected samples included influenza viral RNA concentrations ranging from 10^2^ to 10^5^ gene copies/L (**Tables S7-S9**). In total we selected 60, 58, and 69 wastewater samples positive for InfA H1, InfA H3, and InfB, respectively. More specifically, we selected 15, 15, 29, and 1 InfA H1 wastewater samples at 10^2^, 10^3^, 10^4^ and 10^5^ copies/L, respectively; 14, 17, 26, and 1 InfA H3 wastewater samples at 10^2^, 10^3^, 10^4^ and 10^5^ copies/L, respectively; and 16, 28, 24, and 1 InfB wastewater samples at 10^2^, 10^3^, 10^4^ and 10^5^ copies/L, respectively. These samples were sequenced using their respective tiled amplicon primer scheme as described below.

### Viral concentration and nucleic acid extraction

All samples were processed using a combination of Nanotrap^Ⓡ^ Microbiome A Particles and Nanotrap^Ⓡ^ Enhancement Reagent 1 (CERES nanoscience) for sample concentration and the Applied Biosystems MagMAX^TM^ wastewater Ultra DNA/RNA extraction kit (FisherScientific) aboard a KingFisherFlex system (FisherScientific). Briefly, for each wastewater sample processed, 40mL of wastewater were spiked with 3.5 ul of a 10^5 to 10^6 copies/ul Murine Hepatitis Virus (MHV) surrogate whole virus recovery control, ATCC VR-764. Concentration of each new lot of the control was confirmed using the dPCR protocol described below and an MHV specific assay. Thirty-five mL of MHV spiked water was distributed as 5 ml aliquots into 7 ThermoFisher 24 deep well plates containing 75uL of NanotrapⓇ Microbiome A Particles and 15uL of NanotrapⓇ Enhancement Reagent 1. The plates were placed on the Thermofisher KingFisher Flex and run on a 35 mL concentration protocol that was requested directly from CeresNanotrap. Briefly, all sample plates were premixed for 35 seconds, then each plate was sequentially mixed for 7 minutes before collecting the Microbiome A Particles and transferring them to 500uL of MagMAX Lysis Buffer (FisherScientific). For extraction, 500ul, 40ul, and 20ul of MagMAX binding solution, MagMAX Proteinase K, and MagMAX binding beads respectively, were added to the wastewater lysate and subsequently mixed. Wash Buffer 1, Wash Buffer 2, and Elution plates containing 1mL of MagMAX wash buffer, 1mL of 80% ETOH, and 250uL of MagMAX elution buffer respectively, per sample well were prepared and placed on the KingFisher Flex. A KingFisher Flex protocol downloaded from ThermoFisher (KF protocol https://assets.thermofisher.com/TFS-Assets/BID/Automation-Scripts/MagMAX_Wastewater_Flex24_V2.bdz) was used for automated extraction after sample concentration with one exception, increasing the elution volume from 100uL to 250 uL. Briefly, the wastewater lysate mixed with binding beads was mixed for 6 minutes at 65 degrees C, nucleic acid bound binding beads were then transferred and mixed with Wash Buffer 1 for 3 minutes, transferred and mixed with the Wash Buffer 2 (80% ETOH) for 2 minutes, beads were dried for 2 minutes, and then transferred to the elution buffer and mixed for 6 minutes at 75 degrees C. The final elution was then stored at 4 degrees C until initial dPCR was performed within 24-48 hours of extraction, and stored at -80 degrees C for long term storage and before sequencing.

### Influenza virus quantification using dPCR and normalization

Digital-PCR (dPCR) was performed using the QIAcuity OneStep Advanced Probe Kit (Qiagen) and a set of custom primer and probes that target the M1 and NS2 gene segment for influenza A and influenza B, respectively, and the HA gene segment for influenza subtypes H1 and H3 (Table S10). The 40uL reaction contained 10uL of extracted RNA template, 0.5uM forward primer, 0.5uM reverse primer, 0.25uM probe, 10uL OneStep Advanced Probe Master Mix (4x), 0.2uL OneStep RT Mix, 5uL Enhancer GC, and 10uL nuclease-free water. The thermocycling conditions were 50°C for 30 minutes, followed by 95°C for 2 minutes, and then 45 cycles of 95°C for 10 seconds followed by 55°C for 30 seconds.. The reactions were performed on a Qiagen QIAcuity^Ⓡ^ 8 dPCR platform and analyzed using the accompanying software suite.

Thresholds were set manually to determine final concentration. The following formula was used to calculate the initial concentration of any given target within a wastewater sample.

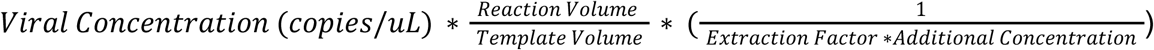

The following formula was used to determine the percent recovery of the MHV wastewater spike in control.

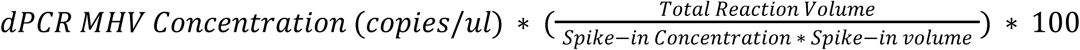

### Tiled Amplicon Primer Design

We designed a tiled amplicon primer scheme to cover the near-complete HA gene segment for seasonal InfA H1, InfA H3 and InfB using Olivar v1.1.12 (*45*). Olivar generates variant-aware tiled amplicon primer schemes by accounting for population level genomic diversity and genomic regions of high mutational frequencies. Primer binding sites are chosen outside regions of high genomic diversity to avoid primer sequence mismatches, which could potentially lead to amplicon dropouts.

To determine the mutational frequency across the HA gene segment required for Olivar, we performed a diversity study of the HA gene segment for InfAH1, InfA H3, and InfB. We downloaded all available HA gene segments of human origin for each virus from January 1, 2018 to April 22, 2023 from the NCBI Influenza Database (https://www.ncbi.nlm.nih.gov/genomes/FLU/Database/nph-select.cgi?go=database). For identical sequences, we selected the sequence with the earliest collection date, discarding the rest. We assigned clade-level classification on all sequences using Nextclade v2.13.1 (*48*). We dropped sequences with unclassified clade assignments and/or if the sequence had less than 85% coverage. This resulted in a total of 4,653, 7,721, and 2,759 unique HA gene sequences, and 15, 28, and 7 distinct HA gene clades for InfAH1, InfA H3, and InfB, respectively. Next, we aligned each sequence to the respective HA gene sequence of the 2023-2024 influenza virus vaccine (A/Victoria/4897/2022 (H1N1), A/Darwin/9/2021 (H3N2) and B/Austria/1359417/2021) using BWA v0.7.17-r1188 (*49*) and samtools v1.21 (*50*). We then determined nucleotide variants relative to the vaccine strain sequence using ivar v1.3.1 (*44*) with minimum allele frequency set to 0 and minimum read depth set to 0. We calculated the frequency of each nucleotide variant found across the HA gene segment and formatted the output as a csv file according to the input requirement for Olivar. We ran Olivar specifying an amplicon size between 125 and 250 basepairs and a seed size of 100. Bed file coordinates were manually corrected so that primer sequences were correctly aligned to the reference sequence (https://github.com/treangenlab/Olivar/issues/14).

### RT-PCR amplification

To create the final primer pools (pool 1 and pool 2) for each primer scheme, we combined either 10uL or 5uL of each primer from 10uM working stocks (**Tables S4, S5, and S6**). We then performed a one-step RT-PCR reaction using Superscript III One-Step (SSIII) RT-PCR System with Platinum Taq High Fidelity DNA Polymerase (Thermo Fisher Scientific). To the reaction mix, we added 0.25 uL of RNase inhibitor, 6uL of pooled primer, and 3uL of extracted nucleic acid for a total reaction volume of 25uL. We ran the reactions on the Eppendorf Mastercyler X50 using the manufacturer’s recommended cycling conditions for <u>SuperScript III One-Step RT-PCR</u> <u>System with Platinum Taq High Fidelity DNA Polymerase</u>, which consisted of cycling at 55℃ for 30 minutes, 94℃ for 2 minutes, followed by 40 cycles of 94℃ for 15 seconds, 60℃ for 30 seconds, 68℃ for 1 minute, a final cycle of 68℃ for 5 minutes, and an infinite hold at 4℃ . After amplification, we combined the products of pool 1 and pool 2 and performed a 1:1 ratio bead cleanup using Illumina purification beads. We quantified nucleic acids using the Qubit^TM^High-Sensitivity dsDNA Quantification Assay Kit (ThermoFisher Scientific) and confirmed amplification of the expected amplicon sizes using the QIAxcel Connect (Qiagen).

### Illumina library preparation and sequencing

We prepared libraries using the Illumina DNA Prep Kit following the Illumina DNA Prep Manufacturer Protocol with minor modifications (https://support-docs.illumina.com/LP/IlluminaDNAPrep/Content/LP/Illumina_DNA/DNA-Prep/Protocol_IDP.htm). Specifically, we increased PCR amplification of tagmented DNA to 7 cycles. We quantified the pooled libraries using a Qubit^TM^ High-Sensitivity dsDNA kit (Thermo Fisher Scientific). We calculated molarity at 1000 base pairs for a 2 nM library and then further diluted to a final loading concentration of 750 pM. The final library pool was loaded into the NextSeq 1000/2000 P1 XLEAP-SBS (300 cycles) cartridge per manufacturers’ instructions and sequenced using the Nextseq 2000 platform to generate 2x150bp reads.

### Clinical sample selection and processing

To assess the potential for amplicon dropouts and/or reduced amplicon coverage that might be explained by reduced primer affinity caused by mismatches between the primer sequence and the currently circulating seasonal influenza sequence diversity, we sequenced 16 clinical samples each of InfA H1, InfA H3 and InfB collected as part of Colorado’s clinical sentinel surveillance system from January 2025 to March 2025 with ct values below 29. Clinical samples were screened using CDC’s Human Influenza Virus Real-Time RT-PCR Diagnostic Panel. Briefly, total nucleic acids were extracted and purified on the KingFisher Flex using the Applied Biosystems MagMAX Viral/Pathogen II Nucleic Acid Isolation Kit (ThermoFisher) and the CDC Emergency Use Only (EUA) Approved FDA Influenza SARS-CoV-2, Influenza A, and Influenza B PCR Assay per the manufacturers instructions (*51,52*, *53*). RT-qPCR was performed using the ABI 7500 Fast Dx. Clinical samples were then sequenced using the same tiled amplicon scheme and following the same protocol as described above for wastewater samples.

### Use of positive controls to evaluate wastewater inhibition

For positive controls, we used whole virus controls in the CDC Influenza Performance Evaluation Program (PEP) Sequencing Panel (Cat # FR-818) purchased from the International Reagent Resource (IRR). From the panel, we selected the following strains to use as controls: A/Idaho/07/2018 (H1N1), A/Hong Kong/45/2019 (H3N2), and B/Colorado/06/2017. At the time of print, this panel has been replaced with the CDC Influenza Next-Gen Sequencing Proficiency Panel (Cat # FR-1794). Total nucleic acids were extracted following the protocol used for clinical samples as described above.

To assess the potential of inhibition caused by wastewater, we performed an 8-step 10-fold serial dilution of the total nucleic acid controls in both a background of nuclease free water (NFW) and RNA extracted wastewater, which had previously tested negative for influenza A and influenza B virus by dPCR (n =1 for each dilution for each background; Table S1, S2, and S3). Control material was sequenced using the same tiled amplicon scheme and following the same protocol as described for wastewater samples.

### Reference guided assembly of the HA gene segment

We used Illumina’s ‘bcl2fastq’ software to convert raw data files in binary base call (BCL) format to FASTQ files and to perform demultiplexing and adapter trimming. We performed additional read filtering and quality trimming using fastp v0.32.2 (*54*). We removed reads that were less than 70 basepairs in length and trimmed reads from the 3’ end with a mean quality score of less than 30 using a 4 basepair sliding window. We determined read quality metrics using Fastqc v0.12.1 (*55*).

We generated bam alignment files and consensus sequences to assess sequencing depth and percent coverages. First, we used BWA v0.7.17-r1188 (*49*) to map quality filtered reads to the HA gene sequence of the 2023-2024 influenza virus vaccine strain (A/Victoria/4897/2022 (H1N1), A/Darwin/9/2021 (H3N2) and B/Austria/1359417/2021). Next, we trimmed primer sequences on aligned reads using samtools amplicon clip v1.21 (*50*) using soft clipping and allowing clipping from both ends. We generated a consensus sequence using ivar v1.3.1 (*44*). For generating the consensus sequence, we specified a base quality score of 20, a minimum read depth of 10, and an allele frequency of 0. We calculated average read depth and percent coverage of the HA gene segment.

To further investigate the specificity of each primer set to their respective subtypes, we performed reciprocal alignments. We performed a full factorial of reciprocal alignments by mapping reads generated with one subtype’s primer set to each of the two other subtype reference genomes (e.g. we mapped reads generated using the Influenza A H1N1 primer set to both the Influenza A H3N2 and Influenza B reference genomes). We manually examined bam files to check for mapped reads and used samtools v1.21 to calculate the mean depth of coverage.

### Single Nucleotide Variant (SNV) Calling

For wastewater samples, which represent a mixed sample of influenza viruses, we performed variant calling to investigate the frequency of nucleotide variants observed across the HA gene segment relative to the HA gene sequences of the 2023-2024 influenza virus vaccine strains (A/Victoria/4897/2022 (H1N1), A/Darwin/9/2021 (H3N2) and B/Austria/1359417/2021). We called nucleotide variants at positions with at least 10x depth and that were present at a frequency of at least 0.03 using iVar variants v. 1.3.1 (*44*).

## Data Availability

The sequencing reads generated from both clinical and wastewater samples were uploaded to NCBI's SRA database and can be accessed under the NCBI BioProject accession PRJNA1293439. All human sequences have been removed from the sequencing reads.
Reference sequences and bed files required for performing reference based alignment and primer trimming can be accessed at https://github.com/CDPHE-bioinformatics/CDPHE-Seasonal-Influenza-Tiled-Amplicon-Sequencing/tree/main.

https://github.com/CDPHE-bioinformatics/CDPHE-Seasonal-Influenza-Tiled-Amplicon-Sequencing/tree/mai

## Acknowledgements

We thank Alexis Alford, Daniel Mallal, and Kevin Castro Vital on the CDPHE Genomic Sequencing team for their contributions in the early development stage and their helpful discussions. We thank Finn Cassidy and Dillon Donaghy on the CDPHE Molecular team for performing clinical sample subtyping. We thank Samuel Baird on the CDPHE Bioinformatics and Genomic Analysis team for helpful discussions on workflow development. We thank the Colorado Wastewater Surveillance Program for their utility coordination and communication efforts.

## Author contributions

Molly C. Hetherington-Rauth and Van Nguyen are designated co-first authors.

Conceptualization: M.C.H-R., V.N., G.L., N.P., L.A.B., A.R., S.R.M.; Investigation: V.N., G.L., N.P..; Data Analysis: M.C.H-R.; Visualizations: M.C.H-R.; Funding Acquisition: L.A.B., A.R., S.R.M.; Supervision: L.A.B., A.R., S.R.M.; Writing -original draft: M.C.H-R.; Writing - review and editing: M.C.H-R., V.N., G.L., N.P., L.A.B., A.R., S.R.M.

## Data and materials availability

The sequencing reads generated from both clinical and wastewater samples were uploaded to NCBI’s SRA database and can be accessed under the NCBI BioProject accession PRJNA1293439. All human sequences have been removed from the sequencing reads.

Reference sequences and bed files required for performing reference based alignment and primer trimming can be accessed at https://github.com/CDPHE-bioinformatics/CDPHE-Seasonal-Influenza-Tiled-Amplicon-Sequencing/tree/main.

## Supplemental Material

**Fig. S1.**
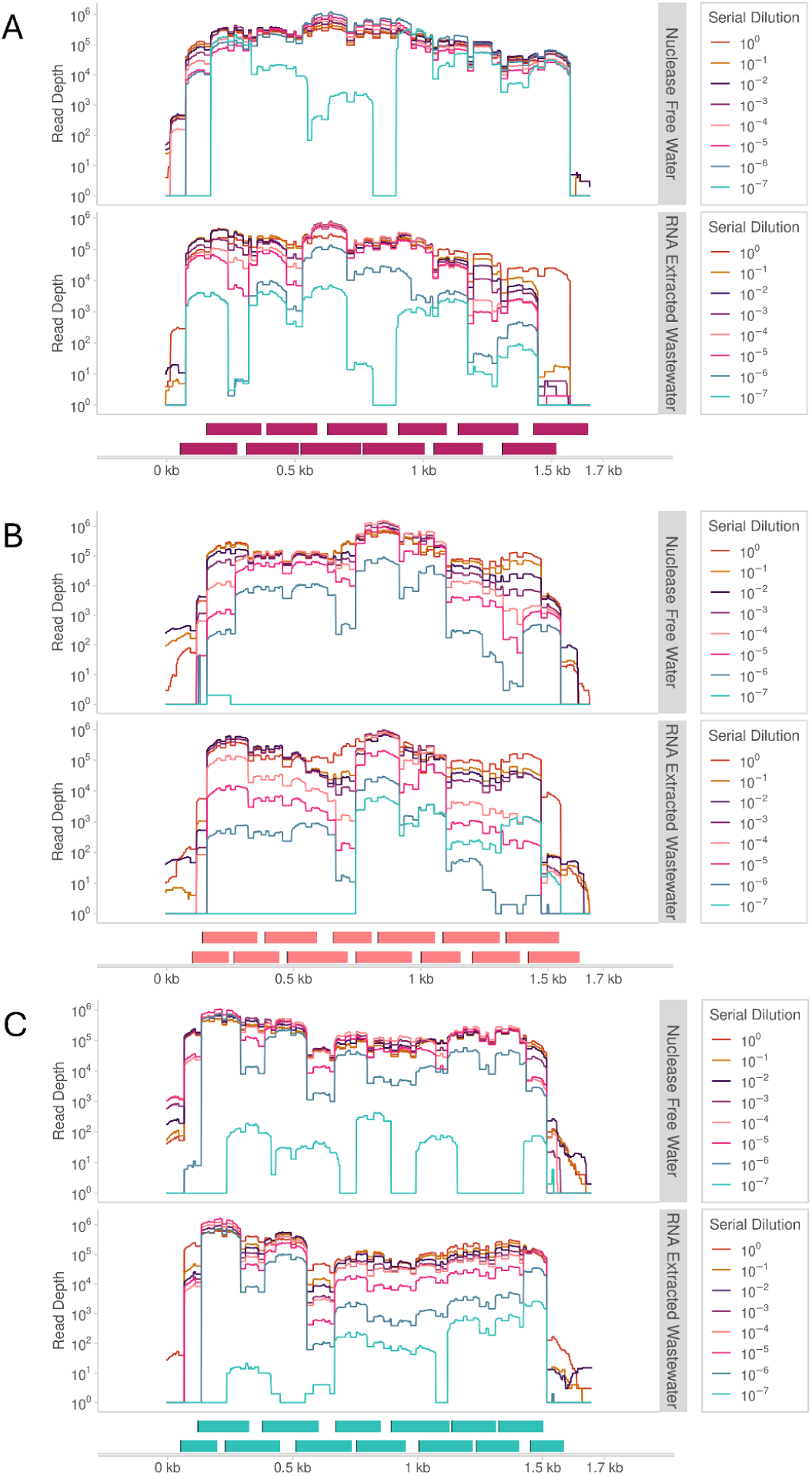
Depth of coverage across the HA gene segment for known whole virus control. We tested our tiled primer scheme on known whole virus controls in a background of either nuclease free water (top track of each panel) or pooled RNA extracted wastewater, which had tested negative for influenza A and B virus by digital PCR (dPCR) (bottom track of each panel) for each (A) InfA H1, (B) InfA H3, and (C) InfB. The following whole virus control strains were used for InfA H1, InfA H3 and InfB, respectively: A/Idaho/07/2018 (H1N1), A/Hong Kong/45/2019 (H3N2), B/Colorado/06/2017. Total nucleic acid was extracted from each whole virus control to a known working concentration. We then performed an 8 step 10-fold serial dilution (10^0^ -10^-7^) on the extracted total nucleic acid (n=1 for each dilution for each background).

**Table S1.**
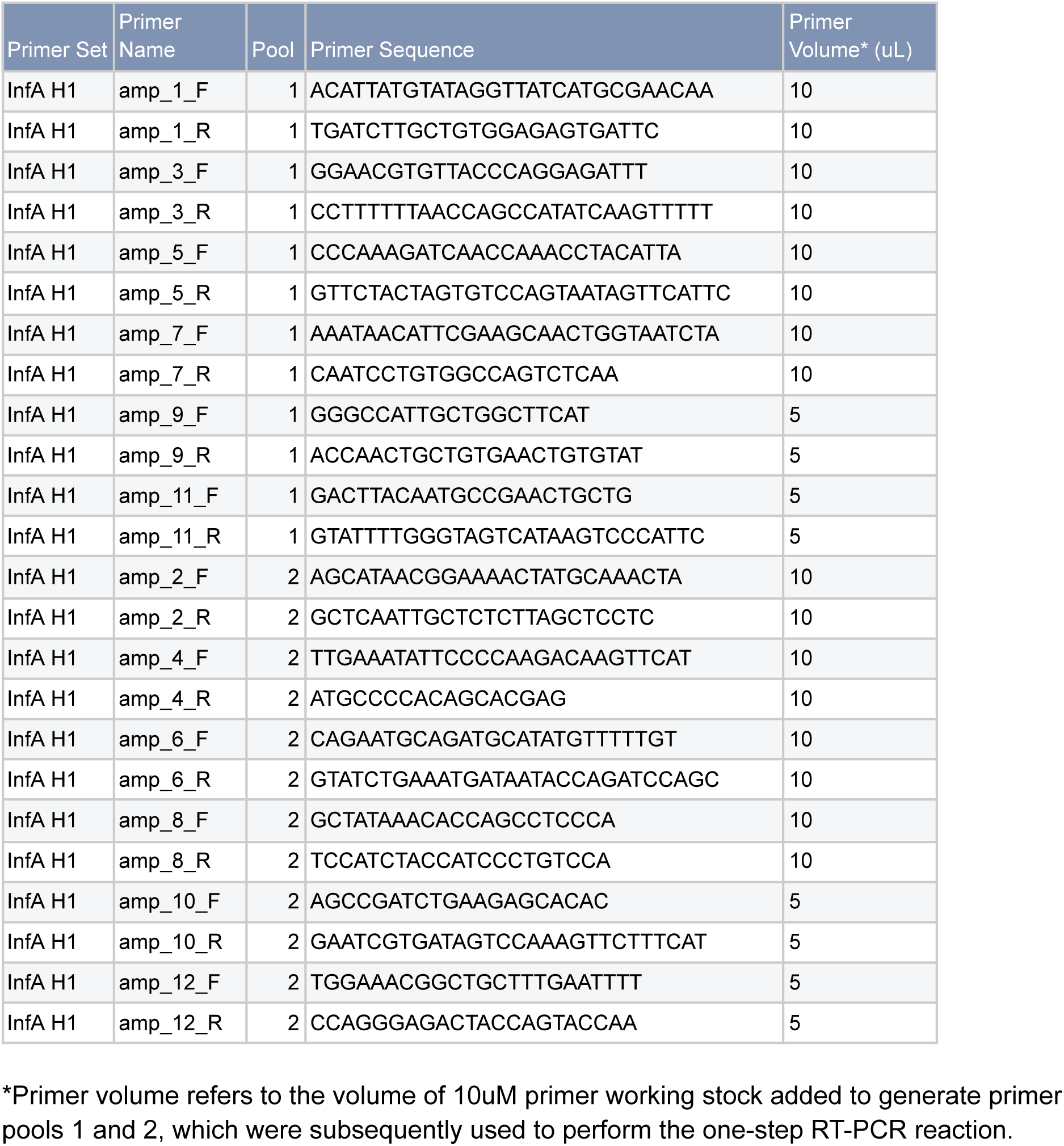
Primer sequences for the InfA H1 primer scheme.

**Table S2.**
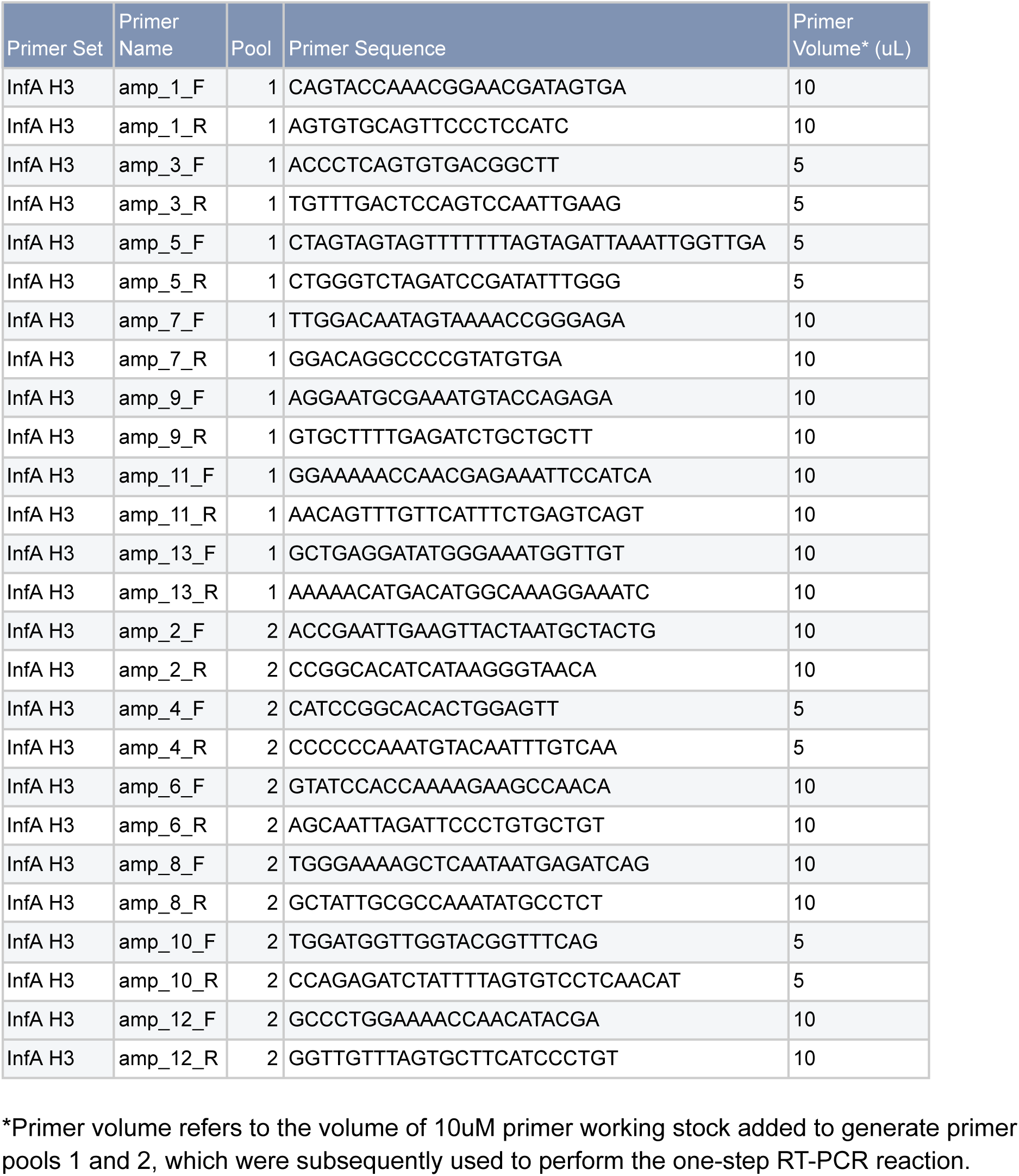
Primer sequences for the InfA H3 primer scheme.

**Table S3.**
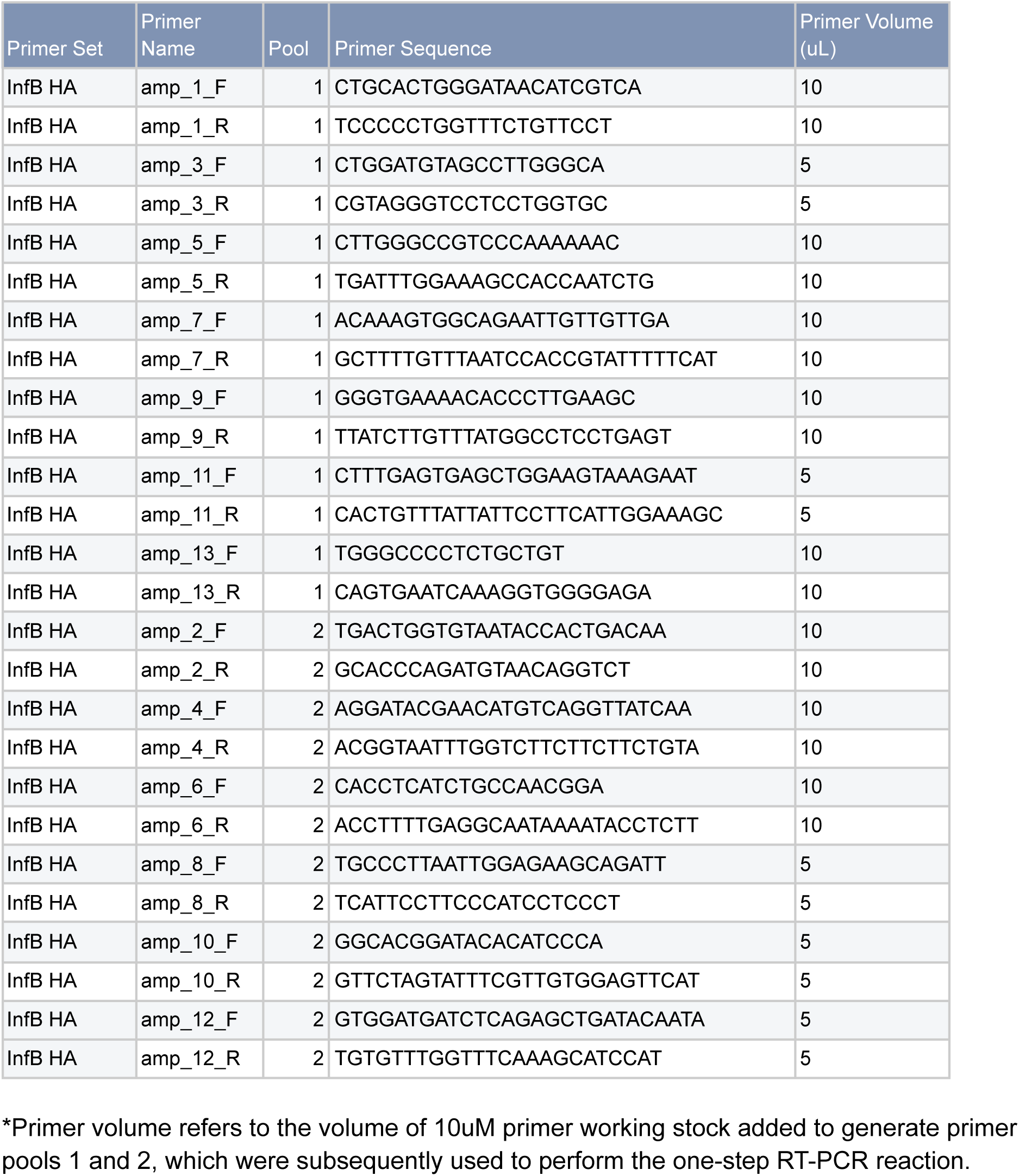
Primer sequences for the InfB primer scheme.

**Table S4.** Assembly metrics of influenza A H1 whole virus control following an eight step, 10-fold serial dilution.

| *Influenza Subtype* | *Background* | *Serial Dilution\** | Count (n) | Mean Depth | % Coverage (Primer Scheme)<sup>†</sup> | % Coverage (HA Gene Segment) | Filtered Reads | Reads Mapped | % Reads Mapped |
| --- | --- | --- | --- | --- | --- | --- | --- | --- | --- |
| Influenza A H1 | Nuclease Free Water | 10<sup>0</sup> | 1 | 145,174 | 104 | 94 | 2,411,460 | 2,406,482 | 99.8 |
|  |  | 10<sup>-1</sup> | 1 | 161,152 | 105 | 95 | 2,678,710 | 2,672,701 | 99.8 |
|  |  | 10<sup>-2</sup> | 1 | 186,513 | 105 | 95 | 3,126,094 | 3,119,322 | 99.8 |
|  |  | 10<sup>-3</sup> | 1 | 170,360 | 105 | 95 | 2,856,264 | 2,851,106 | 99.8 |
|  |  | 10<sup>-4</sup> | 1 | 175,766 | 104 | 94 | 2,911,898 | 2,907,849 | 99.9 |
|  |  | 10<sup>-5</sup> | 1 | 192,336 | 100 | 90 | 3,186,576 | 3,182,335 | 99.9 |
|  |  | 10<sup>-6</sup> | 1 | 228,203 | 100 | 90 | 4,131,392 | 4,123,105 | 99.8 |
|  |  | 10<sup>-7</sup> | 1 | 45,044 | 87 | 79 | 955,316 | 905,359 | 94.8 |
|  | Nuclease Free Waster Total |  | 8 | 163,068 | 101 | 92 | 2,782,214 | 2,771,032 | 99.2 |
|  | Wastewater Extract<sup>‡</sup> | 10<sup>0</sup> | 1 | 115,129 | 104 | 94 | 1,903,222 | 1,899,184 | 99.8 |
|  |  | 10<sup>-1</sup> | 1 | 160,063 | 97 | 87 | 2,917,130 | 2,735,567 | 93.8 |
|  |  | 10<sup>-2</sup> | 1 | 144,479 | 95 | 86 | 2,773,600 | 2,483,180 | 89.5 |
|  |  | 10<sup>-3</sup> | 1 | 134,120 | 92 | 83 | 2,627,162 | 2,292,714 | 87.3 |
|  |  | 10<sup>-4</sup> | 1 | 116,219 | 92 | 83 | 2,444,632 | 1,975,342 | 80.8 |
|  |  | 10<sup>-5</sup> | 1 | 81,098 | 92 | 83 | 2,036,676 | 1,403,375 | 68.9 |
|  |  | 10<sup>-6</sup> | 1 | 14,265 | 86 | 78 | 1,293,880 | 255,642 | 19.8 |
|  |  | 10<sup>-7</sup> | 1 | 1,375 | 73 | 66 | 931,314 | 25,779 | 2.8 |
|  | Wastewater Extract Total |  | 8 | 95,843 | 91 | 82 | 2,115,952 | 1,633,848 | 67.8 |
|  | Total |  | 16 | 129,456 | 96 | 87 | 2,449,083 | 2,202,440 | 83.5 |

\*The working stock concentration was determined by dPCR as 3.29 x 10<sup>6</sup> copies/L.
<sup>†</sup>The primer scheme for InfA H1 90.3% of the HA gene segment.
<sup>‡</sup>Wastewater extract refers to RNA extracted pooled wastewater that tested negative for influenza A and influenza B by dPCR.

**Table S5.** Assembly metrics of influenza A H3 whole virus control following an eight step, 10-fold serial dilution.

| Influenza Subtype | Background | Serial Dilution* | Count (n) | Mean Depth | % Coverage (Primer Scheme)<sup>†</sup> | % Coverage (HA Gene Segment) | Filtered Reads | Reads Mapped | % Reads Mapped |
| --- | --- | --- | --- | --- | --- | --- | --- | --- | --- |
| Influenza A H3 | Nuclease Free Water | 10<sup>0</sup> | 1 | 146,937 | 110 | 94 | 2,484,136 | 2,482,541 | 99.9 |
|  |  | 10<sup>-1</sup> | 1 | 153,219 | 113 | 97 | 2,602,676 | 2,600,678 | 99.9 |
|  |  | 10<sup>-2</sup> | 1 | 157,180 | 113 | 97 | 2,680,796 | 2,678,681 | 99.9 |
|  |  | 10<sup>-3</sup> | 1 | 152,873 | 100 | 86 | 2,954,568 | 2,947,223 | 99.8 |
|  |  | 10<sup>-4</sup> | 1 | 159,101 | 100 | 86 | 3,396,782 | 3,380,999 | 99.5 |
|  |  | 10<sup>-5</sup> | 1 | 102,095 | 100 | 86 | 2,037,026 | 1,938,276 | 95.2 |
|  |  | 10<sup>-6</sup> | 1 | 10,436 | 92 | 79 | 240,202 | 211,030 | 87.9 |
|  |  | 10<sup>-7</sup> | 1 | 0 | 0 | 0 | 71,692 | 2 | 0.0 |
|  | Nuclease Free Waster Total |  | 8 | 110,230 | 91 | 78 | 2,058,485 | 2,029,929 | 85.3 |
|  | Wastewater Extract<sup>‡</sup> | 10<sup>0</sup> | 1 | 172,263 | 113 | 97 | 2,938,524 | 2,936,476 | 99.9 |
|  |  | 10<sup>-1</sup> | 1 | 152,487 | 106 | 91 | 2,693,582 | 2,626,488 | 97.5 |
|  |  | 10<sup>-2</sup> | 1 | 145,205 | 115 | 98 | 2,587,742 | 2,498,219 | 96.5 |
|  |  | 10<sup>-3</sup> | 1 | 158,530 | 100 | 86 | 2,954,794 | 2,745,724 | 92.9 |
|  |  | 10<sup>-4</sup> | 1 | 87,498 | 98 | 84 | 2,506,730 | 1,611,709 | 64.3 |
|  |  | 10<sup>-5</sup> | 1 | 20,414 | 92 | 79 | 1,673,068 | 392,685 | 23.5 |
|  |  | 10<sup>-6</sup> | 1 | 2,461 | 74 | 63 | 1,389,668 | 47,114 | 3.4 |
|  |  | 10<sup>-7</sup> | 1 | 795 | 55 | 47 | 1,275,020 | 15,802 | 1.2 |
|  | Wastewater Extract Total |  | 8 | 92,457 | 94 | 81 | 2,252,391 | 1,609,277 | 59.9 |
|  | Total |  | 16 | 101,343 | 93 | 79 | 2,155,438 | 1,819,603 | 72.6 |

\*The working stock concentration was determined by dPCR as 2.07 x 10<sup>5</sup> copies/L.
<sup>†</sup>The primer scheme for InfA H3 86.4% of the HA gene segment.
<sup>‡</sup> Wastewater extract refers to RNA extracted pooled wastewater that tested negative for influenza A and influenza B by dPCR.

**Table S6.**
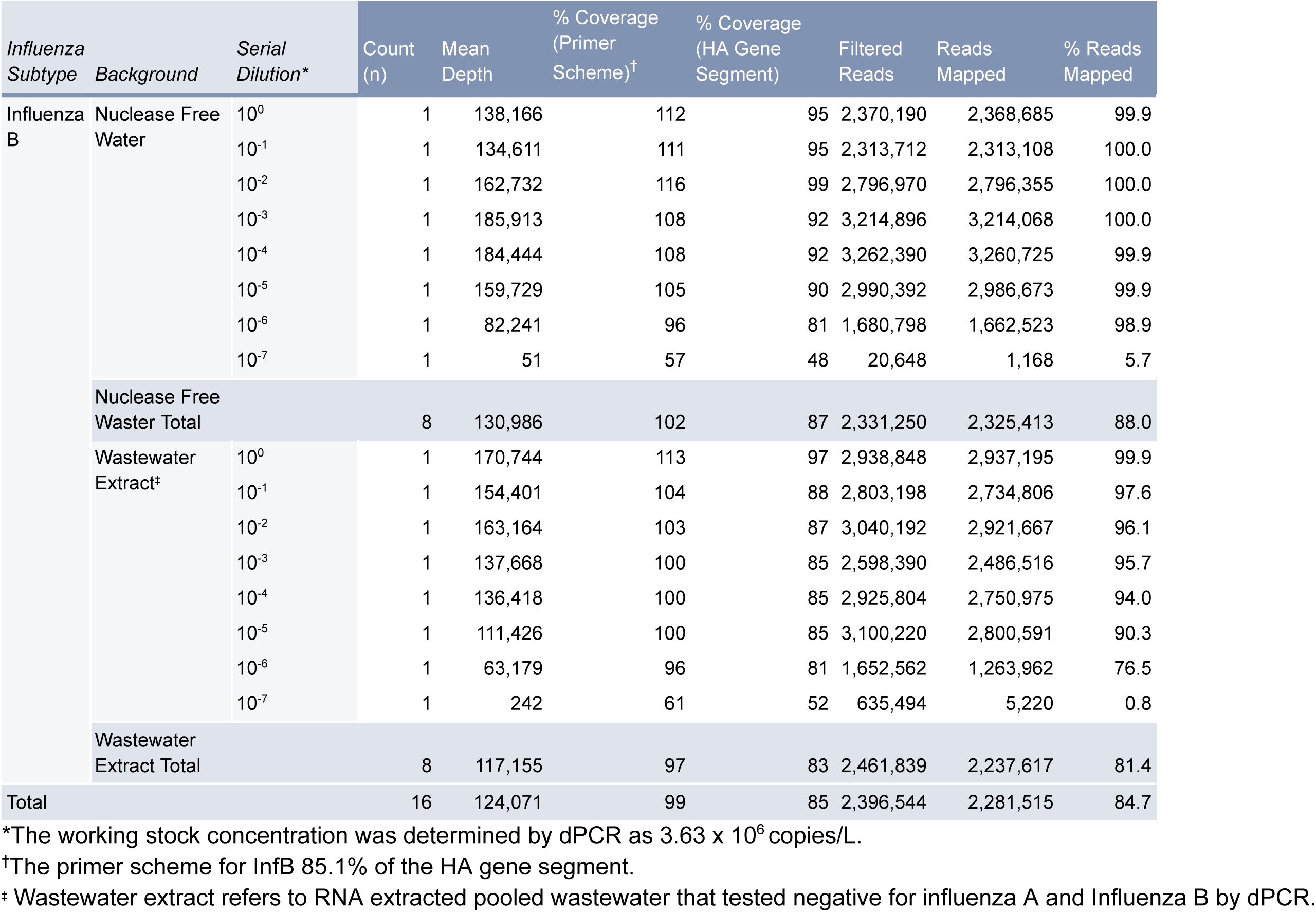
Assembly metrics of Influenza B whole virus control following an eight step, 10-fold serial dilution.

**Table S7.** Wastewater Target Concentrations of Wastewater Samples Sequenced with InfA H1 primer set.

| *Primer Scheme* | Sample Name | InfA Target Concentration (Copies/L) | InfB Target Concentration (Copies/L) | InfA H1 Target Concentration (Copies/L) | InfA H3 Target Concentration (Copies/L) | Collection Date |
| --- | --- | --- | --- | --- | --- | --- |
| InfA H1 | FW-001-AH1 | 7,010 | 629 | 5,490 | 2,190 | 7/24/2024 |
|  | FW-016-AH1 | 0 | 0 | 549 | 549 | 10/17/2024 |
|  | FW-017-AH1 | 2,190 | 1,650 | 549 | 549 | 10/21/2024 |
|  | FW-018-AH1 | 0 | 549 | 549 | 0 | 10/22/2024 |
|  | FW-019-AH1 | 3,300 | 0 | 549 | 9,860 | 10/28/2024 |
|  | FW-020-AH1 | 549 | 0 | 549 | 549 | 10/30/2024 |
|  | FW-021-AH1 | 0 | 0 | 549 | 0 | 10/30/2024 |
|  | FW-022-AH1 | 1,100 | 0 | 549 | 1,650 | 10/30/2024 |
|  | FW-023-AH1 | 16,500 | 0 | 549 | 1,100 | 10/31/2024 |
|  | FW-024-AH1 | 0 | 0 | 549 | 549 | 11/4/2024 |
|  | FW-025-AH1 | 0 | 0 | 549 | 1,100 | 11/18/2024 |
|  | FW-027-AH1 | 1,110 | 0 | 549 | 549 | 11/29/2024 |
|  | FW-031-AH1 | 4,910 | 549 | 2,190 | 6,020 | 12/5/2024 |
|  | FW-032-AH1 | 549 | 560 | 549 | 0 | 12/5/2024 |
|  | FW-033-AH1 | 1,100 | 0 | 549 | 0 | 12/5/2024 |
|  | FW-035-AH1 | 1,100 | 0 | 549 | 1,670 | 12/8/2024 |
|  | FW-037-AH1 | 2,190 | 549 | 3,840 | 0 | 12/9/2024 |
|  | FW-038-AH1 | 0 | 549 | 560 | 1,100 | 12/9/2024 |
|  | FW-042-AH1 | 8,750 | 549 | 13,200 | 7,130 | 12/12/2024 |
|  | FW-043-AH1 | 11,000 | 0 | 5,580 | 7,230 | 12/12/2024 |
|  | FW-051-AH1 | 56,700 | 0 | 13,500 | 10,100 | 12/25/2024 |
|  | FW-053-AH1 | 47,700 | 0 | 9,900 | 4,390 | 12/26/2024 |
|  | FW-054-AH1 | 24,200 | 0 | 37,400 | 54,000 | 12/26/2024 |
|  | FW-055-AH1 | 54,100 | 1,100 | 20,300 | 5,490 | 12/26/2024 |
|  | FW-056-AH1 | 19,300 | 0 | 6,610 | 18,200 | 12/26/2024 |
|  | FW-057-AH1 | 19,100 | 0 | 4,400 | 6,030 | 12/27/2024 |
|  | FW-058-AH1 | 23,600 | 549 | 7,300 | 12,400 | 12/29/2024 |
|  | FW-059-AH1 | 17,500 | 549 | 15,400 | 9,340 | 12/30/2024 |
|  | FW-061-AH1 | 28,000 | 0 | 5,050 | 20,200 | 12/30/2024 |
|  | FW-063-AH1 | 22,000 | 549 | 6,590 | 6,070 | 12/30/2024 |
|  | FW-064-AH1 | 26,300 | 0 | 7,220 | 7,790 | 12/30/2024 |
|  | FW-065-AH1 | 49,000 | 1,100 | 49,000 | 14,900 | 1/1/2025 |
|  | FW-066-AH1 | 12,100 | 0 | 9,910 | 7,150 | 1/2/2025 |
|  | FW-067-AH1 | 22,200 | 0 | 4,240 | 0 | 1/2/2025 |
|  | FW-070-AH1 | 15,000 | 0 | 18,500 | 5,030 | 1/2/2025 |
|  | FW-071-AH1 | 23,800 | 549 | 15,400 | 3,300 | 1/2/2025 |
|  | FW-073-AH1 | 16,400 | 1,100 | 19,300 | 13,100 | 1/2/2025 |
|  | FW-075-AH1 | 116,000 | 606 | 65,900 | 17,700 | 1/3/2025 |
|  | FW-076-AH1 | 5,460 | 617 | 1,980 | 15,200 | 1/3/2025 |
|  | FW-077-AH1 | 39,700 | 0 | 19,600 | 10,400 | 1/6/2025 |
|  | FW-078-AH1 | 11,600 | 0 | 12,900 | 5,380 | 1/6/2025 |
|  | FW-080-AH1 | 606 | 0 | 9,140 | 1,830 | 1/6/2025 |
|  | FW-084-AH1 | 8,250 | 1,660 | 19,800 | 13,200 | 1/9/2025 |
|  | FW-085-AH1 | 38,500 | 1,650 | 51,100 | 36,200 | 1/9/2025 |
|  | FW-087-AH1 | 32,400 | 0 | 46,500 | 13,100 | 1/9/2025 |
|  | FW-098-AH1 | 23,000 | 606 | 21,700 | 14,200 | 1/15/2025 |
|  | FW-100-AH1 | 39,600 | 0 | 19,900 | 12,500 | 1/16/2025 |
|  | FW-101-AH1 | 45,200 | 1,210 | 23,500 | 17,300 | 1/16/2025 |
|  | FW-102-AH1 | 26,900 | 606 | 16,500 | 12,900 | 1/16/2025 |
|  | FW-103-AH1 | 31,800 | 606 | 33,100 | 4,900 | 1/16/2025 |
|  | FW-104-AH1 | 29,900 | 0 | 11,200 | 18,600 | 1/16/2025 |
|  | FW-105-AH1 | 150,000 | 5,520 | 118,000 | 31,600 | 1/16/2025 |
|  | FW-106-AH1 | 65,700 | 0 | 24,600 | 24,600 | 1/16/2025 |
|  | FW-107-AH1 | 64,800 | 1,210 | 49,200 | 15,200 | 1/16/2025 |
|  | FW-108-AH1 | 26,300 | 0 | 26,500 | 13,300 | 1/16/2025 |
|  | FW-109-AH1 | 41,000 | 0 | 23,200 | 29,400 | 1/16/2025 |
|  | FW-110-AH1 | 40,900 | 1,260 | 29,100 | 15,200 | 1/16/2025 |
|  | FW-111-AH1 | 34,900 | 0 | 23,800 | 15,800 | 1/16/2025 |
|  | FW-112-AH1 | 85,300 | 629 | 31,100 | 48,900 | 1/16/2025 |
|  | FW-113-AH1 | 41,900 | 4,890 | 27,300 | 19,300 | 1/16/2025 |

**Table S8.** Wastewater Target Concentrations of Wastewater Samples Sequenced with InfA H3 primer set.

| *Primer Scheme* | Sample Name | InfA Target Concentration (Copies/L) | InfB Target Concentration (Copies/L) | InfA H1 Target Concentration (Copies/L) | InfA H3 Target Concentration (Copies/L) | Collection Date |
| --- | --- | --- | --- | --- | --- | --- |
| InfA H3 | FW-002-AH3 | 2,190 | 549 | 0 | 640 | 8/7/2024 |
|  | FW-003-AH3 | 1,100 | 549 | 0 | 617 | 8/8/2024 |
|  | FW-004-AH3 | 5,460 | 0 | 0 | 594 | 8/15/2024 |
|  | FW-005-AH3 | 2,180 | 0 | 1,260 | 617 | 8/15/2024 |
|  | FW-006-AH3 | 2,190 | 0 | 0 | 674 | 8/15/2024 |
|  | FW-008-AH3 | 560 | 0 | 0 | 629 | 9/23/2024 |
|  | FW-009-AH3 | 0 | 549 | 0 | 663 | 9/26/2024 |
|  | FW-010-AH3 | 549 | 549 | 0 | 617 | 9/26/2024 |
|  | FW-011-AH3 | 0 | 549 | 0 | 629 | 9/26/2024 |
|  | FW-012-AH3 | 2,740 | 2,740 | 0 | 617 | 9/26/2024 |
|  | FW-013-AH3 | 1,650 | 0 | 1,280 | 640 | 10/8/2024 |
|  | FW-026-AH3 | 4,940 | 0 | 549 | 3,370 | 11/21/2024 |
|  | FW-028-AH3 | 13,700 | 549 | 0 | 8,740 | 12/2/2024 |
|  | FW-029-AH3 | 3,840 | 549 | 549 | 1,100 | 12/4/2024 |
|  | FW-030-AH3 | 0 | 560 | 0 | 1,650 | 12/4/2024 |
|  | FW-034-AH3 | 1,670 | 571 | 0 | 560 | 12/5/2024 |
|  | FW-036-AH3 | 11,000 | 549 | 1,100 | 6,580 | 12/9/2024 |
|  | FW-039-AH3 | 7,120 | 549 | 1,100 | 9,290 | 12/9/2024 |
|  | FW-040-AH3 | 1,100 | 549 | 0 | 1,100 | 12/10/2024 |
|  | FW-041-AH3 | 8,790 | 1,100 | 6,640 | 6,630 | 12/12/2024 |
|  | FW-044-AH3 | 28,500 | 549 | 0 | 3,290 | 12/16/2024 |
|  | FW-045-AH3 | 32,900 | 549 | 3,290 | 21,400 | 12/16/2024 |
|  | FW-046-AH3 | 3,290 | 1,650 | 549 | 1,100 | 12/17/2024 |
|  | FW-047-AH3 | 2,730 | 0 | 549 | 1,650 | 12/19/2024 |
|  | FW-049-AH3 | 1,110 | 560 | 0 | 571 | 12/19/2024 |
|  | FW-050-AH3 | 132,000 | 7,700 | 3,840 | 61,900 | 12/20/2024 |
|  | FW-050-AH3 | 132,000 | 7,700 | 3,840 | 61,900 | 12/20/2024 |
|  | FW-052-AH3 | 44,300 | 0 | 25,300 | 21,500 | 12/26/2024 |
|  | FW-054-AH3 | 24,200 | 0 | 37,400 | 54,000 | 12/26/2024 |
|  | FW-060-AH3 | 51,000 | 0 | 23,200 | 13,800 | 12/30/2024 |
|  | FW-062-AH3 | 20,800 | 0 | 7,110 | 9,300 | 12/30/2024 |
|  | FW-063-AH3 | 22,000 | 549 | 6,590 | 6,070 | 12/30/2024 |
|  | FW-068-AH3 | 35,900 | 549 | 15,000 | 27,800 | 1/2/2025 |
|  | FW-069-AH3 | 11,600 | 0 | 7,710 | 8,270 | 1/2/2025 |
|  | FW-072-AH3 | 5,470 | 1,100 | 7,770 | 3,700 | 1/2/2025 |
|  | FW-074-AH3 | 109,000 | 0 | 1,120 | 59,900 | 1/2/2025 |
|  | FW-079-AH3 | 9,650 | 606 | 15,700 | 13,900 | 1/6/2025 |
|  | FW-081-AH3 | 6,100 | 606 | 6,720 | 8,870 | 1/6/2025 |
|  | FW-082-AH3 | 54,400 | 3,850 | 28,100 | 25,100 | 1/7/2025 |
|  | FW-083-AH3 | 12,600 | 0 | 11,500 | 3,830 | 1/9/2025 |
|  | FW-084-AH3 | 8,250 | 1,660 | 19,800 | 13,200 | 1/9/2025 |
|  | FW-086-AH3 | 36,600 | 0 | 12,200 | 17,000 | 1/9/2025 |
|  | FW-088-AH3 | 13,200 | 0 | 22,000 | 13,700 | 1/9/2025 |
|  | FW-089-AH3 | 13,200 | 1,650 | 14,400 | 13,900 | 1/9/2025 |
|  | FW-091-AH3 | 38,100 | 0 | 11,000 | 13,400 | 1/13/2025 |
|  | FW-092-AH3 | 42,200 | 5,930 | 16,700 | 10,100 | 1/13/2025 |
|  | FW-093-AH3 | 47,500 | 0 | 31,200 | 13,600 | 1/13/2025 |
|  | FW-094-AH3 | 143,000 | 1,310 | 96,900 | 26,200 | 1/13/2025 |
|  | FW-095-AH3 | 15,500 | 629 | 11,600 | 22,700 | 1/14/2025 |
|  | FW-096-AH3 | 20,400 | 1,250 | 14,400 | 24,000 | 1/14/2025 |
|  | FW-097-AH3 | 35,200 | 1,780 | 14,800 | 12,900 | 1/14/2025 |
|  | FW-099-AH3 | 32,700 | 1,820 | 2,500 | 16,300 | 1/15/2025 |
|  | FW-116-AH3 | 19,100 | 0 | 7,660 | 11,500 | 1/21/2025 |
|  | FW-118-AH3 | 34,000 | 2,740 | 16,100 | 12,200 | 1/23/2025 |
|  | FW-120-AH3 | 96,900 | 0 | 5,590 | 160,000 | 1/28/2025 |
|  | FW-122-AH3 | 88,800 | 2,440 | 549 | 36,700 | 1/30/2025 |
|  | FW-123-AH3 | 32,400 | 0 | 7,130 | 615 | 1/30/2025 |
|  | FW-124-AH3 | 8,200 | 631 | 1,270 | 635 | 1/30/2025 |
|  | FW-125-AH3 | 65,200 | 553 | 1,100 | 48,900 | 1/31/2025 |

**Table S9.** Wastewater Target Concentrations of Wastewater Samples Sequenced with InfB primer set.

| *Primer Scheme* | Sample Name | InfA Target Concentration (Copies/L) | InfB Target Concentration (Copies/L) | InfA H1 Target Concentration (Copies/L) | InfA H3 Target Concentration (Copies/L) | Collection Date |
| --- | --- | --- | --- | --- | --- | --- |
| InfB | FW-007-BHA | 0 | 640 |  |  | 9/9/2024 |
|  | FW-014-BHA | 0 | 57,600 | 0 | 0 | 10/10/2024 |
|  | FW-015-BHA | 0 | 19,200 | 0 | 0 | 10/10/2024 |
|  | FW-048-BHA | 20,800 | 6,010 | 9,350 | 7,750 | 12/19/2024 |
|  | FW-090-BHA | 71,200 | 20,900 | 20,400 | 30,000 | 1/10/2025 |
|  | FW-114-BHA | 56,500 | 76,900 | 40,700 | 17,600 | 1/17/2025 |
|  | FW-115-BHA | 123,000 | 25,700 | 25,700 | 26,800 | 1/21/2025 |
|  | FW-117-BHA | 142,000 | 10,900 | 39,000 | 47,200 | 1/23/2025 |
|  | FW-119-BHA | 46,000 | 7,660 | 8,760 | 8,780 | 1/27/2025 |
|  | FW-121-BHA | 110,000 | 32,200 | 4,800 | 13,200 | 1/30/2025 |
|  | FW-126-BHA | 45,400 | 16,400 | 22,100 | 10,100 | 2/3/2025 |
|  | FW-127-BHA | 18,300 | 820 | 8,790 | 0 | 2/6/2025 |
|  | FW-128-BHA | 120,800 | 62,080 | 132,600 | 9,017 | 2/13/2025 |
|  | FW-129-BHA | 40,800 | 24,600 | 37,300 | 602 | 2/20/2025 |
|  | FW-130-BHA | 3,280 | 12,200 | 6,090 | 0 | 2/24/2025 |
|  | FW-131-BHA | 36,200 | 3,830 | 1,650 | 1,100 | 2/26/2025 |
|  | FW-132-BHA | 38,000 | 40,800 | 17,000 | 1,120 | 2/27/2025 |
|  | FW-133-BHA | 20,300 | 7,120 | 18,700 | 6,300 | 2/27/2025 |
|  | FW-134-BHA | 34,500 | 1,090 | 2,200 | 4,960 | 2/27/2025 |
|  | FW-135-BHA | 16,500 | 550 | 1,100 | 550 | 2/27/2025 |
|  | FW-136-BHA | 12,600 | 546 | 1,100 | 1,100 | 2/27/2025 |
|  | FW-137-BHA | 23,100 | 554 | 6,070 | 556 | 2/27/2025 |
|  | FW-138-BHA | 27,500 | 14,300 | 11,000 | 2,200 | 2/27/2025 |
|  | FW-139-BHA | 9,850 | 547 | 1,100 | 1,100 | 2/27/2025 |
|  | FW-140-BHA | 40,100 | 1,100 | 3,830 | 1,100 | 2/27/2025 |
|  | FW-141-BHA | 24,100 | 1,730 | 9,910 | 1,100 | 2/27/2025 |
|  | FW-142-BHA | 71,800 | 5,030 | 0 | 549 | 2/28/2025 |
|  | FW-143-BHA | 3,830 | 547 | 609 | 613 | 3/2/2025 |
|  | FW-144-BHA | 22,200 | 1,840 | 0 | 0 | 3/3/2025 |
|  | FW-145-BHA | 13,800 | 26,400 | 0 | 0 | 3/3/2025 |
|  | FW-146-BHA | 122,000 | 0 | 3,840 | 547 | 3/3/2025 |
|  | FW-147-BHA | 9,300 | 7,110 | 14,300 | 5,590 | 3/3/2025 |
|  | FW-148-BHA | 13,700 | 1,640 | 1,210 | 3,620 | 3/3/2025 |
|  | FW-149-BHA | 10,100 | 18,400 | 603 | 1,210 | 3/4/2025 |
|  | FW-150-BHA | 21,900 | 147,000 | 1,110 | 4,980 | 3/6/2025 |
|  | FW-151-BHA | 12,100 | 551 | 0 | 3,290 | 3/6/2025 |
|  | FW-152-BHA | 33,000 | 552 | 4,380 | 3,290 | 3/6/2025 |
|  | FW-153-BHA | 10,400 | 1,090 | 0 | 1,650 | 3/6/2025 |
|  | FW-154-BHA | 57,400 | 2,750 | 547 | 8,750 | 3/6/2025 |
|  | FW-155-BHA | 20,800 | 1,090 | 1,090 | 7,650 | 3/6/2025 |
|  | FW-156-BHA | 31,800 | 19,700 | 2,740 | 9,310 | 3/6/2025 |
|  | FW-157-BHA | 13,500 | 1,100 | 1,100 | 1,650 | 3/6/2025 |
|  | FW-158-BHA | 27,500 | 1,650 | 0 | 2,200 | 3/6/2025 |
|  | FW-159-BHA | 13,900 | 1,100 | 1,100 | 3,840 | 3/6/2025 |
|  | FW-160-BHA | 4,590 | 1,750 | 1,100 | 547 | 3/6/2025 |
|  | FW-161-BHA | 8,770 | 548 | 549 | 1,100 | 3/6/2025 |
|  | FW-162-BHA | 18,600 | 1,640 |  |  | 3/6/2025 |
|  | FW-163-BHA | 23,600 | 1,100 | 0 | 9,940 | 3/6/2025 |
|  | FW-164-BHA | 13,200 | 1,100 | 1,100 | 4,380 | 3/6/2025 |
|  | FW-165-BHA | 5,510 | 17,000 | 0 | 552 | 3/7/2025 |
|  | FW-166-BHA | 548 | 0 | 9,300 | 0 | 3/7/2025 |
|  | FW-167-BHA | 2,190 | 548 | 548 | 0 | 3/10/2025 |
|  | FW-168-BHA | 6,190 | 0 | 0 | 0 | 3/10/2025 |
|  | FW-169-BHA | 7,640 | 5,460 | 1,100 | 2,740 | 3/10/2025 |
|  | FW-170-BHA | 2,740 | 547 | 547 | 1,100 | 3/10/2025 |
|  | FW-171-BHA | 4,380 | 1,090 | 0 | 1,100 | 3/10/2025 |
|  | FW-172-BHA | 1,640 | 547 | 0 | 548 | 3/10/2025 |
|  | FW-173-BHA | 1,100 | 1,640 | 0 | 1,100 | 3/10/2025 |
|  | FW-174-BHA | 15,900 | 7,660 | 3,840 | 1,650 | 3/10/2025 |
|  | FW-175-BHA | 5,480 | 1,110 | 1,650 | 0 | 3/10/2025 |
|  | FW-176-BHA | 1,650 | 549 | 0 | 1,110 | 3/10/2025 |
|  | FW-177-BHA | 4,400 | 29,700 | 557 | 0 | 3/10/2025 |
|  | FW-178-BHA | 1,690 | 561 | 0 | 0 | 3/11/2025 |
|  | FW-179-BHA | 1,780 | 13,100 | 2,400 | 1,800 | 3/13/2025 |
|  | FW-180-BHA | 4,950 | 11,600 | 0 | 1,110 | 3/24/2025 |
|  | FW-181-BHA | 14,300 | 14,900 | 0 | 550 | 3/24/2025 |
|  | FW-182-BHA | 6,570 | 21,500 | 1,110 | 1,670 | 3/25/2025 |
|  | FW-183-BHA | 13,700 | 55,400 | 5,480 | 1,100 | 3/26/2025 |
|  | FW-184-BHA | 595 | 28,100 | 546 | 0 | 3/28/2025 |

**Table S10.**
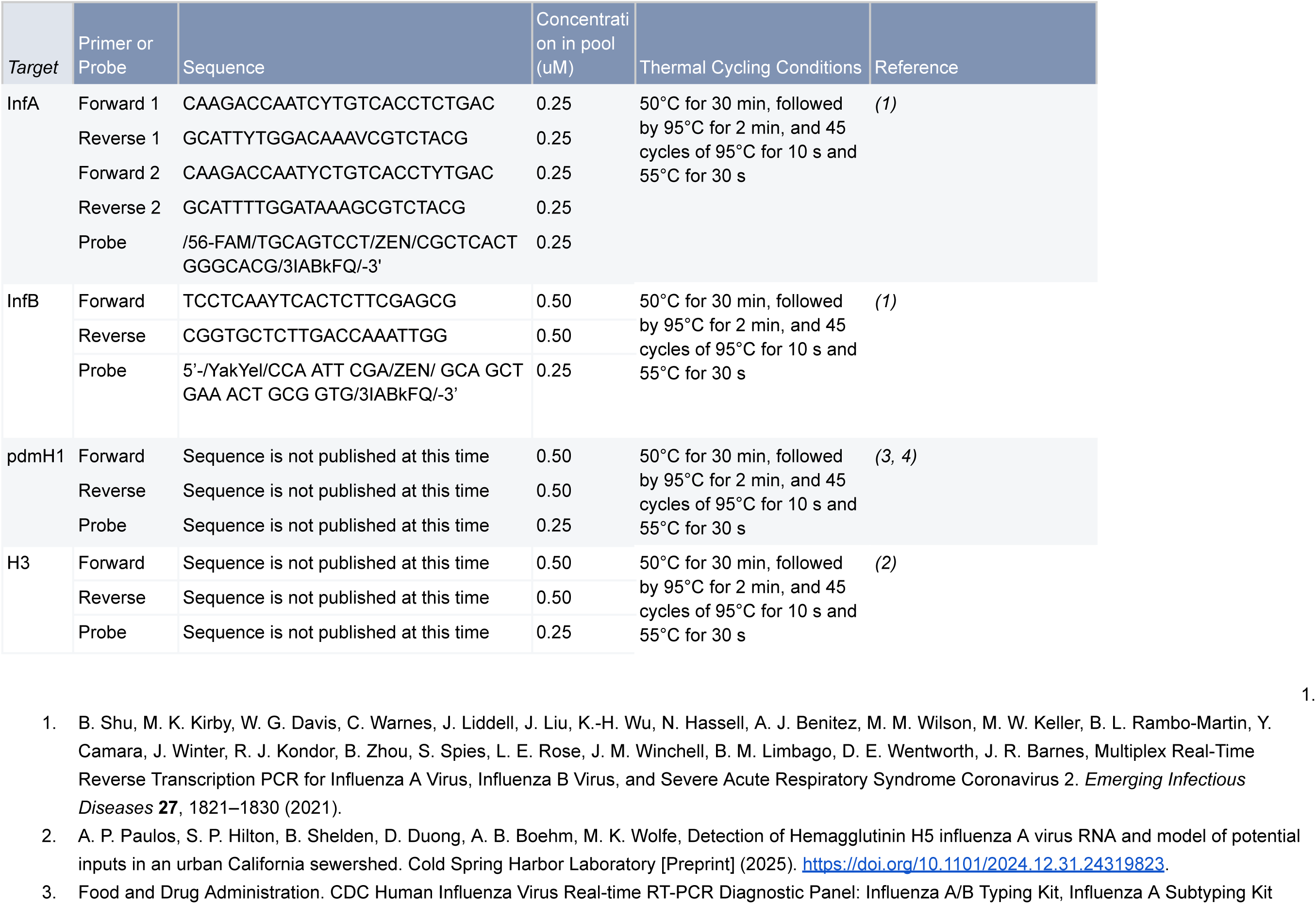
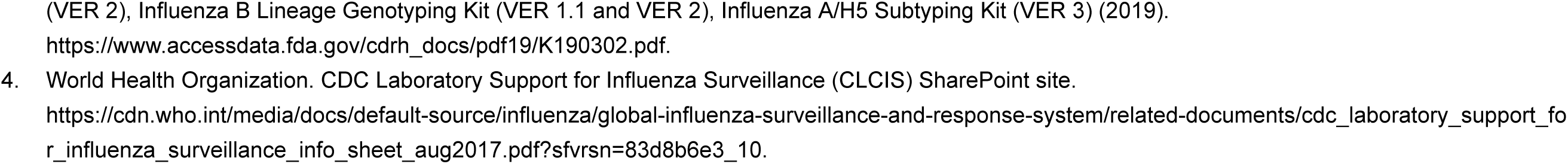
dPCR primer and probe sequences. pdmH1 (3,4) and H3 (2) subtyping primers are unpublished at this time.

